# Integrative multi-omics approaches identify molecular pathways and improve Alzheimer’s Disease risk prediction

**DOI:** 10.1101/2025.05.31.25328688

**Authors:** Rasika Venkatesh, Katie M. Cardone, Yuki Bradford, Anni K. Moore, Rachit Kumar, Jason H. Moore, Li Shen, Dokyoon Kim, Marylyn D. Ritchie

## Abstract

Alzheimer’s Disease (AD) is the most prevalent condition that impacts the aging population, with no effective treatment or singular underlying causal factor identified. As a complex disease, characterizing the genetic risk of developing AD has proven to be difficult; polygenic scores (PGS) exclusively use common variants which fail to fully capture disease heterogeneity. This study used univariate and multivariate approaches to characterize AD risk. Genome-, transcriptome-, and proteome-wide association studies (GWAS, TWAS, and PWAS) were conducted on 15,480 individuals from the Alzheimer’s Disease Sequencing Project (ADSP) R4 release to identify AD-associated signals, followed by pathway enrichment analysis. Integrative risk models (IRMs) were developed using genetically-regulated components of gene and protein expression and clinical covariates. Elastic-net logistic regression and random forest classifiers were evaluated using area under the receiver operating characteristic (AUROC), area under the precision-recall curve (AUPRC), F1-score, and balanced accuracy. These IRMs were compared against baseline PGS and covariate models. We identified 104 genomic, 319 transcriptomic, and 17 proteomic associations with AD under significant thresholds. Putatively novel associations were enriched in signaling, myeloid differentiation, and immune pathways. The best-performing IRM, random forest with transcriptomic and covariate features, achieved an AUROC of 0.703 and AUPRC of 0.622, significantly outperforming PGS and baseline models. Integrating univariate discovery approaches with multivariate modeling enhances AD risk prediction and offers insights into underlying biological processes.

## 1. Background

Alzheimer’s disease (AD) is the most prevalent neurodegenerative condition affecting the aging population, posing a significant public health challenge as life expectancy continues to rise[1,2]. Late-onset AD (LOAD), the most common form of AD, has proven particularly challenging to characterize in terms of genetic risk[1]. Much of the genetic contribution to LOAD remains unexplained, complicating efforts to develop predictive models[2]. Most existing models rely on genetic variants identified through genome-wide association studies (GWAS). The APOE ε4 allele is the most well-established genetic risk factor for AD, accounting for one-quarter of the heritable contributions to liability, with total heritability of AD estimated to be between 58 - 75%[3–5]. The contributions of variants in other genes, such as *BIN1*, have also been characterized via large-scale GWAS[4,6].

Emerging evidence indicates that AD-associated genetic variants converge on key biological pathways. These include cholesterol and lipid metabolism, neuroinflammation, and synaptic function[7]. Disruptions in these processes contribute to hallmark AD pathologies such as amyloid-β accumulation, tau hyperphosphorylation, and neuronal loss[7]. This biological context has spurred a shift toward approaches that use transcriptomic, proteomic, and clinical data with genomic information to uncover the molecular mechanisms underlying AD. Recent evidence from other complex traits suggest that tissue-specific gene expression and protein expression derived from transcriptome- and proteome-wide association studies (TWAS and PWAS) provide orthogonal and complementary information to genetic data alone, offering opportunities to improve risk prediction models and identify putatively novel genes and proteins[8,9]. TWAS have identified 54 hippocampal genes linked to AD risk, with fine-mapping prioritizing 24 candidates (e.g., *PICALM*, *BIN1*) whose effects are mediated through tissue-specific expression[10]. Proteome-wide analyses reveal 43 AD-associated proteins, including *TOMM40* and *APOC1*, validated through mediation testing of pQTL effects (63% concordance)[11]. These associations were detected due to increased statistical power and reduced multiple testing burden, suggesting that these approaches can aid in identifying the underlying mechanisms of complex traits[10,11].

Polygenic scores (PGS), which aggregate the effects of multiple common genetic variants identified from GWAS, have historically underperformed in predicting AD risk, as AD is influenced by a combination of genetic, molecular, and environmental factors. The limitations of PGS models for AD prediction are well-documented[3,12–14]. Leonenko *et al* reported that PGS for AD computed using PRS-CS achieved area under the receiver operating characteristic (AUROC) values ranging from 0.55 to 0.75[12]. Their best-performing model, incorporating genome-wide significant single nucleotide polymorphisms (SNPs) and *APOE*, achieved an R^2^ of 0.24. Notably, excluding *APOE* from the model reduced the R^2^ to 0.06, highlighting the influence of *APOE* on AD risk prediction[12]. Instead, models that incorporate functional information may provide deeper insights into disease etiology and offer more robust tools for individualized risk assessment. Studies developing gene expression risk scores (GeRS) with PGS demonstrated improved predictive performance across phenotypes, underscoring the potential of multimodal approaches in enhancing risk prediction[15]. Furthermore, ensemble machine learning approaches in PGS for SNP selection and modeling have been shown to better account for non-linear and interaction effects, suggesting that incorporating nonlinear modeling techniques with multi-modal data could further enhance prediction accuracy[16].

This study explores the use of univariate and multivariate approaches, paired with multi-modal data, to improve risk prediction for LOAD in one of the most comprehensive AD datasets currently available. We use univariate approaches—GWAS, TWAS, and PWAS—to identify associations within each molecular layer. These approaches are valuable for detecting individual signals but do not capture interactions across modalities. To capture complementary biological information beyond genetic data alone, we use multivariate modeling to develop Integrative Risk Models (IRMs) that integrate transcriptomic and proteomic features (Figure 1), enhancing predictive accuracy and enabling a more comprehensive understanding of disease risk. By addressing the limitations of current approaches and exploring the added value of multi-modal molecular data, we aim to advance understanding of AD risk and improve predictive accuracy for clinical and research applications.

**Figure 1.**
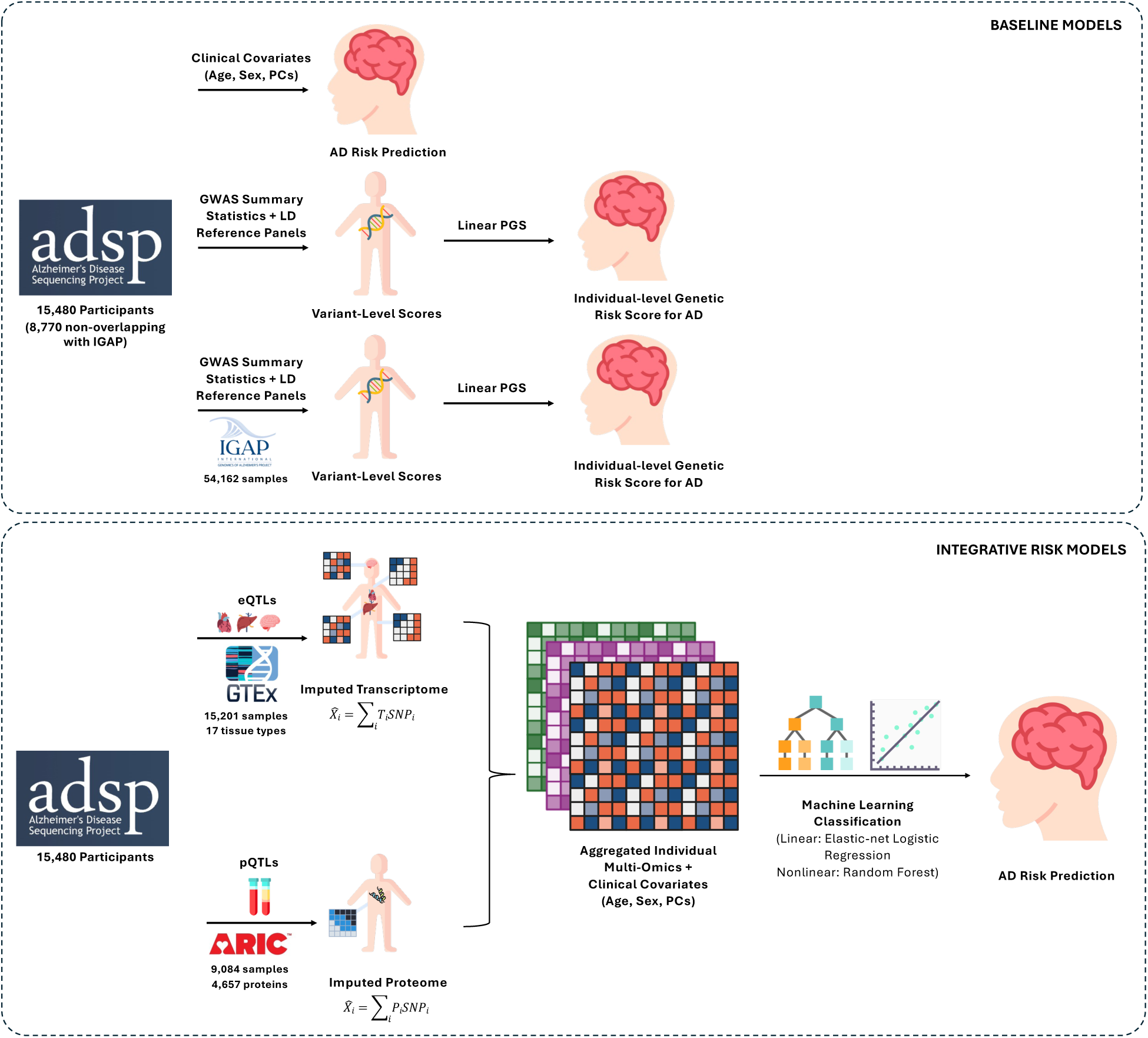
Overview of IRM framework. 15,480 participants from ADSP, eQTLs from GTEx, and pQTLs from ARIC were used to impute gene and protein expression. This information is combined with clinical covariates of age, sex, and PCs and then used as features for machine learning classification algorithms. Baseline models of clinical covariates and 2 separate PGS were generated to evaluate performance against the IRM.

## 2. Methods

### 2.1 Alzheimer’s Disease Sequencing Project (ADSP) Quality Control

The Alzheimer’s Disease Sequencing Project (ADSP) R4 dataset from the National Institute on Aging Genetics of Alzheimer’s Disease Data Storage Site (NIAGADS) includes whole genome sequencing (WGS) data from a globally diverse population of 11,384 individuals diagnosed with AD, 16,685 individuals without AD, and 8,292 individuals with uncertain or alternative diagnoses, spanning 40 global cohorts[17–19]. The full ADSP dataset includes data from nearly all previous large-scale AD studies, including the Alzheimer’s Disease Neuroimaging Initiative (ADNI), Religious Orders Study and Memory and Aging Project (ROS/MAP), and the Anti-Amyloid Treatment in Asymptomatic Alzheimer’s Disease (A4 Study), among several other cohorts, thus creating one of the most comprehensive existing datasets for AD and related dementias[20–23]. Resequencing, variant calling, and various quality control (QC) measures were conducted in accordance with the standards established by ADSP, ensuring high-quality genomic data[24]. Additionally, case and control phenotypes were harmonized across cohorts by ADSP experts to reduce variability in diagnostic criteria and improve comparability prior to data release[17,18,24].

For the present study, the initial dataset of 36,361 individuals was subsetted based on defined criteria to enable focused analysis of late-onset Alzheimer’s disease (LOAD). We selected cohorts with a mean age of cases greater than or equal to 70 years, thereby excluding early-onset cohorts. Additional considerations included ensuring balanced sex distribution among cases and controls, by ensuring at least 30% female cases, removing cohorts with very low case-control counts (percentage of cases < 15%), and excluding samples with mixed phenotypes such as other dementias. The resulting subset comprised 15,480 participants - 6,885 cases and 8,595 controls - where cases were determined by the harmonized diagnosis score being > 4. Principal component analysis (PCA) was performed to assess the genetic similarity of this global population to labeled reference populations by projecting the ADSP participants against the 1000 Genomes reference populations[7]. The full participant counts by cohort after subsetting and QC are available in **Supplemental Table S1**.

### 2.2 Genome-wide Association Study (GWAS)

We conducted a genome-wide association study (GWAS) using PLINK v2.0 after performing subsetting and performing QC on the ADSP dataset to perform an initial analysis to discern genetic loci associated with AD risk[25]. As part of QC measures prior to performing GWAS, variant names were standardized by filling in missing identifiers. Variants that failed laboratory-based QC filters were removed, along with intentionally duplicated samples. Additional filtering steps included removing variants with a minor allele count (MAC) less than 20, applying a 99% variant call rate threshold, and a 95% sample call rate threshold. Analyses were conducted both with and without applying a minor allele frequency (MAF) threshold of 0.01 to assess sensitivity to rare variant inclusion[26]. To ensure robustness and account for potential confounders, our models were adjusted for various parameters, including age at diagnosis for cases or age at date of data release for controls, sex, and the first 5 principal components (PCs) to account for population stratification[26]. Significant loci were identified at a genome-wide significant p-value threshold of p < 5E-08.

### 2.3 Transcriptome-wide Association Study (TWAS)

To study tissue-specific gene-expression changes associated with AD, we conducted transcriptome-wide association studies (TWASs) using PrediXcan and multivariate adaptive shrinkage (MASHR) eQTL models from the Genotype-Tissue Expression (GTEx) Project v8, available in PredictDB[27,28]. GTEx eQTLs were derived from a sample group consisting of 84.6% European ancestry, 12.9% African American, 1.3% Asian and 1.1% unknown ancestry, a multi-ancestry cohort similar to that of ADSP[29]. Using this reference, we imputed genetically regulated gene expression (GReX) for all protein coding genes in 17 tissues known to be relevant in Alzheimer’s Disease and neurodegenerative disorders (brain spinal cord cervical c-1, brain cortex, brain substantia nigra, colon sigmoid, liver, vagina, whole blood, brain anterior cingulate cortex BA24, brain frontal cortex BA9, brain hypothalamus, brain caudate basal ganglia, brain nucleus accumbens basal ganglia, brain cerebellar hemisphere, brain putamen basal ganglia, brain cerebellum, brain amygdala, and brain hippocampus)[28–32]. Associations with AD status were calculated independently for each of these 12 tissues. Significant genes were determined using a Bonferroni threshold (p < 0.05/# genes tested) per tissue.

### 2.4 Proteome-wide Association Study (PWAS)

We then performed a proteome-wide association study (PWAS) using PrediXcan with the WGS data for the individuals in the ADSP subset. PWAS identifies genetic associations that may influence complex traits, such as AD, by regulating protein abundance in tissue[33]. Blood plasma-derived protein quantitative trait loci (cis-pQTLs) from the Atherosclerosis Risk in Communities (ARIC) study were used to construct the models[34]. This large bi-ethnic study was made up of 9,084 participants, consisting of 7,213 European Americans (EA) and 1,871 African Americans (AA). PrediXCan PWAS EA and AA models were identified in PredictDB, constructed using ARIC consortium data by utilizing PEERS covariates, expression information from eQTL associations, as well as gene and SNP annotations[34,35]. The genetically-regulated components of protein expression were imputed using the EA and AA cohort information, after which PWAS was performed. The resulting PWAS associations were assessed for statistical significance using a Bonferroni threshold (p < 0.05/# proteins tested) for each model.

### 2.5 Gene-Set Enrichment Analysis

Gene-set enrichment was performed using Enrichr for the significant results from GWAS, TWAS and PWAS, respectively[36–38]. Enrichment analysis explored the specific pathways and processes associated with the statistically significant genes and proteins from each association study. Pathway results were annotated with KEGG 2021, Reactome 2022, and Gene Ontology (GO) Biological Process 2023 pathways[37]. The significant pathways were identified as having a Fisher’s exact test adjusted p-value < 0.05[37].

### 2.6 Baseline Models

#### 2.6.1 Baseline Polygenic Scores (PGS)

A baseline PGS was created and evaluated on the ADSP dataset, calculated using PRS-CS, a Bayesian regression framework that infers posterior SNP effect sizes under continuous shrinkage (CS) priors using GWAS summary statistics and an external linkage disequilibrium (LD) reference panel[39]. We applied PRS-CS to the ADSP data using GWAS summary statistics generated from 10 iterations of split-sample analyses within ADSP, treating each split as the discovery dataset. To ensure robust and unbiased estimation of the PGS, we performed 10 independent iterations of training and testing. In each iteration, the ADSP dataset was randomly split into 80% training and 20% testing subsets. PRS-CS was applied to the training subset using LD information from a multi-ancestry LD reference panel from 1000 Genomes Project individuals, producing posterior SNP effect sizes for PGS computation[40]. Individual-level PGS values were then calculated using the --score function in PLINK 2.0 by summing the dosages of risk alleles weighted by the PRS-CS posterior effect sizes[25]. After obtaining PGS values for each individual, logistic regression models were trained using the PGS as a single predictor for AD status in the 80% training set of each iteration. Model performance was evaluated on the held-out 20% from each iteration, and the final model performance was estimated by averaging metrics such as AUC, sensitivity, and specificity across the 10 test sets. To further assess generalizability, we conducted a nested 80/20 evaluation within the original 20% held-out data from each iteration. This involved splitting the 20% test set into an 80/20 internal validation/test subset, fitting a logistic regression model using the previously computed PGS on the 80%, and evaluating its performance on the internal 20% to simulate model behavior in unseen data. This iterative evaluation approach ensured that the baseline PGS model performance was assessed with reduced risk of overfitting and better reflects the expected predictive power in the ADSP cohort.

A secondary PGS was created using GWAS summary statistics from the International Genomics of Alzheimer’s Disease (IGAP) study Stage 1 exclusively with PRS-CS (n = 54,162)[39,41]. A linkage disequilibrium (LD) reference panel of European ancestry individuals from the 1000 Genomes phase 3 project was utilized in the computation[40]. The AD summary statistics from IGAP are the largest set of summary statistics currently available for this phenotype, providing a robust dataset to use for a baseline PGS. The APOE e4 variant (rs429358) was initially excluded from the computation as it was not on the LD reference panel; this variant was added back as a comparison model, using the IGAP GWAS beta as this variant’s PGS. To avoid overfitting and biased sampling, the PGS was applied using PLINK2’s --score function to individuals in ADSP who do not overlap with IGAP and met phenotyping criteria described above (n = 8,770). The resulting PGS was fitted with a LASSO regression, using AD case/control status as the outcome and age, sex, and PC1-5 as covariates. The sample was split into a 70% and 30% train/test split with 100 random iterations, balanced by case/control ratio. Performance was assessed with average AUROC, area under the precision-recall curve (AUPRC), F1 score and balanced accuracy across the iterations.

#### 2.6.2 Genetic Similarity Ablation Study

We additionally conducted an ablation study to evaluate the impact of population structure on model performance. Two baseline elastic net logistic regression models were constructed to classify risk of AD. The first model included the first 5 PCs derived from genome-wide genetic data, along with age and sex as covariates, to account for genetic similarity and demographic confounding. The second model excluded the PCs, retaining only age and sex as covariates. This comparison allowed us to assess the contribution of genetic population structure to the predictive accuracy and performance of the classification model.

### 2.7 Integrated Risk Models (IRMs)

#### 2.7.1 Elastic Net Logistic Regression Models with Imputed Data Modalities

After QC, the ADSP dataset consisted of 15,480 participants from the subset, including 6,885 cases and 8,595 controls. Participants were selected based on predefined phenotypic criteria, and demographic variables such as age and sex. To account for population structure and minimize confounding effects from population structure, the first 5 principal components (PCs) were also incorporated as features in the analysis.

Between 1,232 - 10,985 imputed gene and protein expression features were used as inputs to each model, with the exact number depending on the tissue type, as determined by the GTEx project, and protein expression model used from the ARIC study. The full set of models with feature counts are available in **Supplemental Table S2**. An elastic net regression model was developed to predict case-control status for each combination of imputed gene expression tissue and imputed protein expression, leveraging a combination of L1 (LASSO) and L2 (ridge) regularization. This approach was utilized due to its ability to handle multicollinearity among predictors while simultaneously performing feature selection and coefficient shrinkage[42]. The dataset was divided into training, validation, and test sets using a 70:15:15 split and scaled column-wise using z-score normalization, by standardizing each feature by removing the mean and scaling to unit variance[43]. The training set was used to fit the model, while the validation set was reserved for hyperparameter tuning. An independent test set was used exclusively for final model evaluation to ensure unbiased performance assessment. Hyperparameter tuning was conducted using grid search with five-fold cross-validation to optimize the L1 ratio, which controls the balance between lasso and ridge penalties, and the regularization strength (C) parameter[42]. The full set of hyperparameters tuned are shown in **Supplemental Table S3**. To ensure model convergence during training, the maximum number of iterations was set to 500. To address the high dimensionality of the dataset, 2 feature selection processes were tested. Least absolute shrinkage and selection operator (LASSO) regression was employed, as well as principal component analysis (PCA), which also reduced the dimensionality of the selected features while retaining the most informative components for downstream modeling[44]. PCA was fit on the scaled training set to extract the minimum number of principal components required to explain 80% of the total variance of the dataset. The same transformation was applied to the validation and test sets to ensure consistency across datasets. Of these tests, PCA was determined to most improve performance by reducing more features while maintaining orthogonality in the data.

The comprehensive train-test-validation split framework allowed for robust hyperparameter optimization while mitigating overfitting risks. The final elastic net models were evaluated on the test set to determine predictive performance. The AUROC, AUPRC, F1-score, and balanced accuracy metrics were used to assess the model’s ability to discriminate between cases and controls. Ten iterations of each model were run, to get measures of the average and standard deviation of each metric. This approach ensured that the model’s performance was robust and generalizable across the diverse tissue-specific datasets analyzed in the study.

#### 2.7.2 Random Forest Models with Imputed Data Modalities

To address the limitations of linear modeling approaches in capturing complex, nonlinear relationships between genetic, transcriptomic, and proteomic features, we implemented a series of random forest models[43]. Random forests are well-suited for exploring nonlinear patterns and interactions within high-dimensional datasets[45]. This approach was applied to the same subset of data from ADSP, with the same covariates of age, sex, and first 5 PCs. The dataset was split into 70:15:15 training, validation, and test sets. The training set was used to fit the random forest model, while the validation set was reserved for hyperparameter tuning. After identifying the optimal hyperparameter combination, the model was evaluated on an independent test set to ensure unbiased performance assessment.

To optimize the random forest models, we employed random search, a strategy that efficiently samples from predefined hyperparameter distributions to identify optimal configurations. This method was chosen for its computational efficiency compared to exhaustive grid search. Hyperparameters of the number of estimators, maximum tree depth, minimum samples required to split a node, minimum samples required at a leaf node, and maximum features considered for a split were all tuned[46]. The full set of hyperparameters tuned are shown in **Supplemental Table S3**. Random search was performed over 10 iterations, with five-fold cross-validation used to evaluate model performance for each hyperparameter combination. This iterative process ensured that the selected model configuration for every iteration of each random forest model was optimal for performance and generalizable.

Model performance was measured using AUROC, AUPRC, F1-score, and balanced accuracy. Ten iterations of each model were run, to get measures of the average and standard deviation of each metric. These metrics provided a robust evaluation of the model’s ability to discriminate between cases and controls. An illustrative overview of the baseline models and IRM framework is depicted in **Figure 1**.

## 3. Results

### 3.1 ADSP Study Population

Following resequencing, variant calling, and quality control processes adhering to ADSP protocols, phenotype harmonization across cohorts ensured consistent diagnostic criteria for subsequent analyses. Principal component analysis (PCA) revealed that 3 to 4 principal components (PCs) accounted for 80% of the explained variance in this subset (**Supplemental Figure S1A**). Furthermore, projecting the PCs onto the 1000 Genomes reference population (**Supplemental Figure S1B)** showed a similar genetic similarity distribution to the full ADSP R4 dataset, suggesting that population-specific information was not lost through the subsetting approach.

After subsetting the dataset based on the exclusion criteria described in Methods and projecting the PCA of the participants on the 1000 Genomes reference populations, the remaining dataset of 15,480 individuals indicates that sex and case-control status were distributed across five major genetic-similarity groups: African (AFR), Admixed American (AMR), European (EUR), South Asian (SAS), and Other (**Table 1**). The majority of participants self-identified or genetically clustered as EUR (5,241 individuals), followed by AMR (5,049), AFR (4,623), Other (546), and SAS (21). Within each population group, both cases and controls were represented across sexes, with minor imbalances observed among the control populations. For example, the AFR subgroup had more female cases (1,172) than male cases (844), and a higher count of male controls (2,157) than female controls (450). Representation from SAS and "Other" groups was limited, comprising fewer than 600 individuals in total, though both case and control statuses were included. Overall, the subsetted dataset retained diversity across population groups, and ensured sex was balanced within most groups, supporting downstream analyses. The full participant counts by cohort are available in **Supplemental Table S1**.

**Table 1.** Participant counts from the ADSP subset, stratified by case-control status, biological sex, and genetically-inferred ancestry.

| **Genetic Similarity Group** | **Sex** | **Case/Control Status** | **Number of Participants** |
| --- | --- | --- | --- |
| AFR | Female | Control | 2157 |
| AFR | Female | Case | 1172 |
| AFR | Male | Control | 844 |
| AFR | Male | Case | 450 |
| AMR | Female | Control | 2188 |
| AMR | Female | Case | 1270 |
| AMR | Male | Control | 957 |
| AMR | Male | Case | 634 |
| EUR | Female | Control | 1322 |
| EUR | Female | Case | 1737 |
| EUR | Male | Control | 850 |
| EUR | Male | Case | 1332 |
| Other | Female | Control | 154 |
| Other | Female | Case | 178 |
| Other | Male | Control | 110 |
| Other | Male | Case | 104 |
| SAS | Female | Control | 7 |
| SAS | Female | Case | 5 |
| SAS | Male | Control | 6 |
| SAS | Male | Case | 3 |

### 3.2 Association Study and Pathway Enrichment Analyses

We identified 104 genomic, 319 transcriptomic, and 76 proteomic associations with AD via GWAS (p-value < 5E-08), TWAS (p-value < 5.62E-06), and PWAS (p < 5.29E-05), respectively under genome-wide significance thresholds for GWAS and Bonferroni-significance thresholds for TWAS and PWAS (**Figure 2A**). Manhattan plots and QQ plots of the GWAS results are available in **Supplemental Figure S2A-C**. The significant GWAS risk loci mapped to 12 unique genes by position, 94 unique genes were identified via TWAS, and 17 unique genes identified via PWAS. Of these associations, 9 genes and proteins, including *APOE*, *APOC1, TOMM40, NECTIN2, WWC1, NRCAM, KCNV2*, *CR1,* and APP were validated in the Alzheimer’s Disease Variant Portal (ADVP)[47]. The remaining associations are putatively novel, though some, such as *BLOC1S5-TXNDC5,* have been linked to AD progression in the literature, but have not been previously reported in the ADVP[47,48]. *GLCE* was implicated across all 3 association studies; this gene is linked to heparan sulfate production and was found in previous studies to be upregulated in AD patients[49]. The full tables of statistically significant associations for each association study are available in **Supplemental Tables S4-6**, and the set of unique annotated genes and proteins and their ADVP status are available in **Supplemental Table S7**.

**Figure 2.**
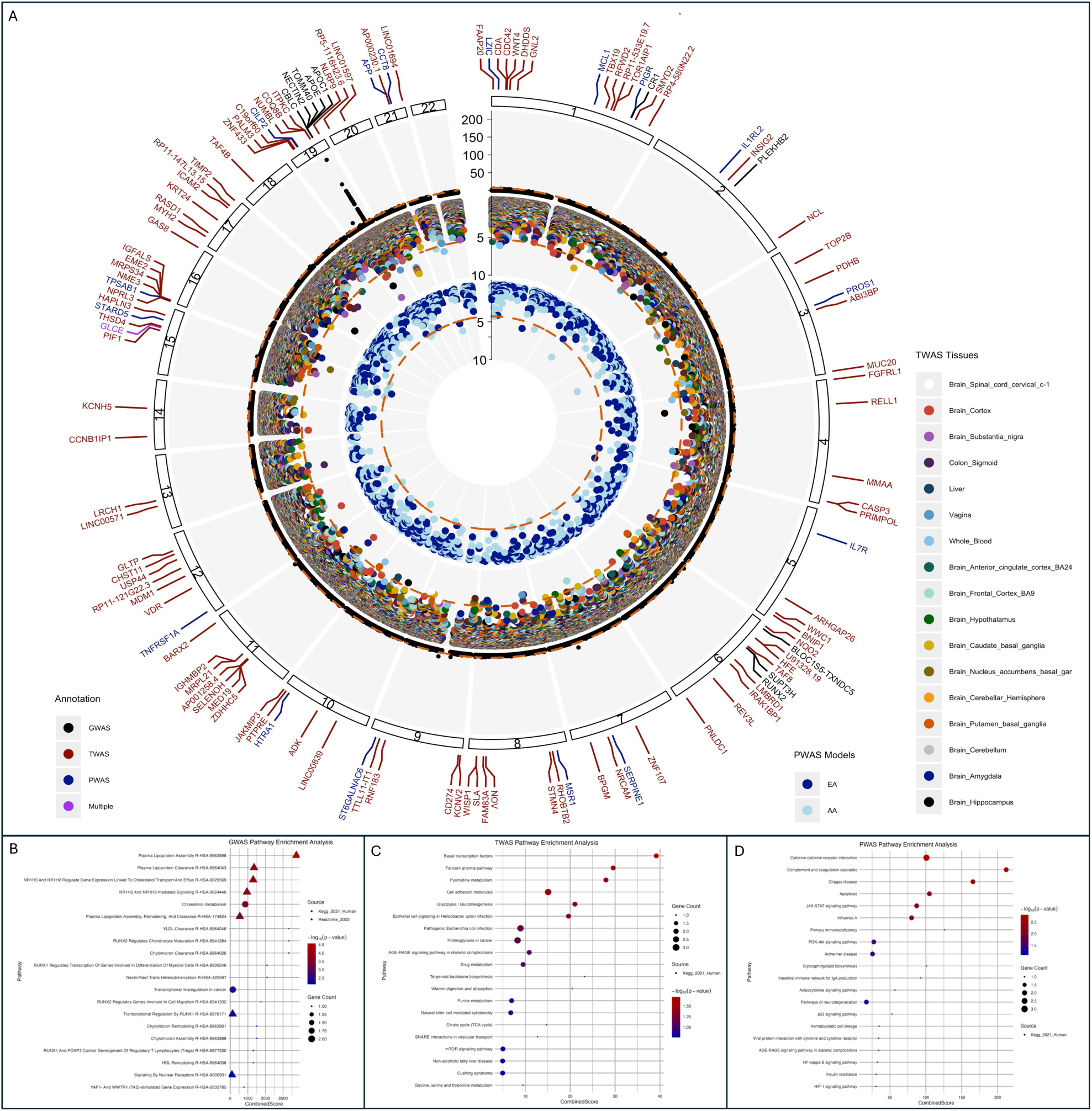
Results of association studies and pathway analyses. (A) Circos plot depicting all association study results by tissue and model type. Significant results appear above the dashed orange line denoting Bonferroni significance thresholds. (B-D) KEGG, Reactome, and GO gene-set enrichment results for GWAS, TWAS and PWAS, colored by p-value. Size of the points denotes the number of gene or protein inputs contributing to the pathway.

In order to identify the known biologically relevant pathways associated with the statistically significant GWAS, TWAS, and PWAS results, gene-set enrichment analysis was performed using EnrichR for Reactome 2022, KEGG 2021, and Gene ontology (GO) 2023 pathways[37]. Significant pathways were identified at an adjusted p-value < 0.05 from the Fisher’s Exact Test; the full pathway results for each association study are available in **Supplemental Table S8**. Pathway analysis of GWAS results (**Figure 2B**) identified significant overrepresentation of lipid metabolism and cholesterol transport pathways, particularly involving *APOC1* and *APOE*. The most enriched pathway was Plasma Lipoprotein Assembly (adjusted p = 2.69E-05), followed by Plasma Lipoprotein Clearance (adjusted p = 1.33E-04) and NR1H3 and NR1H2 Regulate Gene Expression Linked to Cholesterol Transport and Efflux (adjusted p = 1.40E-04). GO terms related to Phospholipid Efflux (adjusted p = 1.23E-05) and High-Density Lipoprotein Particle Remodeling (adjusted p = 2.35E-05) were also significantly enriched. These findings underscore the central role of cholesterol homeostasis in Alzheimer’s disease pathogenesis, consistent with the known involvement of *APOE* in lipid transport and amyloid-beta metabolism[7]. Additional enriched pathways included Cholesterol Efflux (adjusted p = 5.66E-05), and broader processes such as Cholesterol Metabolism (adjusted p = 2.72E-04) and Plasma Lipoprotein Assembly, Remodeling, and Clearance (adjusted p = 5.49E-04), reinforcing the relevance of lipid regulation mechanisms in genetic susceptibility to Alzheimer’s disease[7]. Although no pathways reached statistical significance under the adjusted p-value in the TWAS results, several suggestively significant biological processes in neurodegeneration and genomic maintenance were highly ranked (**Figure 2C**). Among the top-ranked pathways were Error-Prone Translesion Synthesis (adjusted p = 0.2394) and Postreplication Repair (adjusted p = 0.2394), involving genes such as *REV3L* and *PRIMPOL*, which are involved in DNA damage tolerance[50].

The PWAS pathway results revealed several immune-related pathways with statistically significant enrichment, suggesting a potential link between immune dysregulation and AD (**Figure 2D)**. The top-enriched pathway was Signaling by Interleukins (adjusted p = 2.07E-05), driven by proteins including APP, IL1RL2, IL7R, TNFRSF1A, and MCL1, all of which are implicated in inflammatory signaling cascades. The Immune System pathway (adjusted p = 4.92E-05) was also enriched, encompassing proteins such as APP, IL1RL2, PROS1, PIGR, and CCT8. Several related processes including Cytokine Signaling in the Immune System, Regulation of Inflammatory Response (adjusted p = 3.29E-05), and Positive Regulation of Defense Response further support the hypothesis that systemic immune modulation plays a role in Alzheimer’s pathophysiology. Notably, APP, an important contributor to amyloid pathology, appeared consistently across these pathways, underscoring the potential interplay between amyloid biology and immune signaling at the protein level[11].

### 3.3 IRM Model Performance

The performance of the elastic-net logistic regression models demonstrated a progressive improvement with the inclusion of additional feature sets, as seen in **Table 2**. The baseline linear model using only age and sex yielded the lowest performance across all metrics (AUROC = 0.546, AUPRC = 0.459, Balanced Accuracy = 0.496, F1-score = 0.080). The inclusion of PCs as covariates to account for individual-level genetic similarity substantially improved performance (AUROC = 0.636, AUPRC = 0.536, Balanced Accuracy = 0.604, F1-score = 0.521), indicating that population structure and genetic similarity play a significant role in predictive accuracy. The PGS alone achieved AUROC = 0.608 but resulted in a lower AUPRC (0.491) compared to the covariate-only model, suggesting that PGS alone may not capture enough variability in the phenotype. Interestingly, combining PGS with covariates did not yield further improvement beyond the covariate-only model, underscoring the potential limitations of PGS in this context. The PGS conducted using scores derived from IGAP demonstrated some performance improvement in AUROC relative to the PGS performed on splits of ADSP, as expected given the large sample size and robust design of the IGAP study (AUROC = 0.641). However, by the other metrics, this model performed much worse (AUPRC = 0.446, Balanced Accuracy = 0.507, F1-score = 0.0413). These results highlight the discrepancy between AUROC and other relevant performance metrics when evaluating models across populations. Overall performance was lacking compared to the multi-omics models.

**Table 2.** Performance metrics of the baseline models and IRMs - AUROC, AUPRC, Balanced Accuracy, and F1 Score. Values represent the mean performance across iterations with corresponding standard deviations and 95% confidence intervals.

| Model | Features | AUROC | AUPRC | Balanced Accuracy | F1-Score |
| --- | --- | --- | --- | --- | --- |
| Elastic-Net Logistic Regression | Age + Sex | 0.546 (0.006)<br>[0.542, 0.550] | 0.459 (0.007)<br>[0.455, 0.463] | 0.496 (0.004)<br>[0.493, 0.498] | 0.080 (0.028)<br>[0.062, 0.097] |
|  | Age + Sex + PCs (Covariates) | 0.636 (0.007)<br>[0.632, 0.640] | 0.536 (0.008)<br>[0.531, 0.541] | 0.604 (0.007)<br>[0.600, 0.608] | 0.521 (0.01)<br>[0.515, 0.527] |
|  | ADSP PGS (Genome) | 0.608 (0.016)<br>[0.604, 0.611] | 0.491 (0.015)<br>[0.488, 0.494] | 0.540 (0.013)<br>[0.537, 0.542] | **0.691 (0.011)**<br>**[0.689, 0.693]** |
|  | ADSP PGS + Covariates | 0.623 (0.017)<br>[0.620, 0.627] | 0.484 (0.015)<br>[0.481, 0.487] | 0.555 (0.015)<br>[0.552, 0.558] | 0.681 (0.012)<br>[0.679, 0.684] |
|  | IGAP PGS + Covariates | 0.641 (0.001)<br>[0.640, 0.642] | 0.446 (0.001)<br>[0.444, 0.449] | 0.507 (0.001)<br>[0.506, 0.508] | 0.041 (0.002)<br>[0.037, 0.046] |
|  | Transcriptome + Covariates | 0.652 (0.010)<br>[0.649, 0.655] | 0.566 (0.013)<br>[0.563, 0.570] | 0.605 (0.009)<br>[0.603, 0.608] | 0.522 (0.012)<br>[0.518, 0.525] |
|  | Proteome + Covariates | 0.631 (0.008)<br>[0.628, 0.635] | 0.548 (0.011)<br>[0.543, 0.552] | 0.602 (0.007)<br>[0.599, 0.605] | 0.521 (0.010)<br>[0.517, 0.526] |
|  | Transcriptome + Proteome_EA + Covariates | 0.660 (0.014)<br>[0.654, 0.663] | 0.521 (0.020)<br>[0.516, 0.526] | 0.574 (0.009)<br>[0.572, 0.577] | 0.396 (0.014)<br>[0.392, 0.400] |
|  | Transcriptome + Proteome_AA + Covariates | 0.640 (0.009)<br>[0.638, 0.642] | 0.557 (0.013)<br>[0.553, 0.561] | 0.602 (0.008)<br>[0.599, 0.604] | 0.525 (0.014)<br>[0.521, 0.528] |
| Random Forest | Transcriptome + Covariates | **0.703 (0.010)**<br>**[0.699, 0.707]** | **0.622 (0.012)**<br>**[0.617, 0.627]** | **0.645 (0.007)**<br>**[0.642, 0.648]** | 0.616 (0.009)<br>[0.613, 0.620] |
|  | Proteome + Covariates | 0.680 (0.005)<br>[0.677, 0.682] | 0.599 (0.007)<br>[0.595, 0.603] | 0.636 (0.003)<br>[0.634, 0.637] | 0.624 (0.008)<br>[0.620, 0.627] |
|  | Transcriptome + Proteome_EA + Covariates | 0.689 (0.009)<br>[0.685, 0.693] | 0.613 (0.014)<br>[0.607, 0.618] | 0.637 (0.005)<br>[0.635, 0.639] | 0.620 (0.011)<br>[0.615, 0.624] |
|  | Transcriptome + Proteome_AA + Covariates | 0.690 (0.009)<br>[0.686, 0.694] | 0.613 (0.014)<br>[0.607, 0.618] | 0.637 (0.005)<br>[0.635, 0.639] | 0.617 (0.013)<br>[0.612, 0.623] |

The integration of multi-omic data showed notable gains, with the transcriptome + covariates model outperforming PGS-based models (AUROC = 0.652, AUPRC = 0.566, Balanced Accuracy = 0.605). The top 5 tissues based on median performance were selected - brain cortex, whole blood, brain frontal cortex BA9, brain nucleus accumbens basal ganglia, brain substantia nigra - and performance metrics were averaged across model iterations. The proteome + covariates model performed slightly worse (AUROC = 0.631, AUPRC = 0.548), while integrating both transcriptomic and proteomic data (EA and AA models) produced varying results, with the EA model achieving higher AUROC (0.660) but lower AUPRC (0.521), suggesting differences in predictive power.

The random forest models outperformed the logistic regression models across all feature sets, demonstrating the advantage of non-linear methods in capturing complex relationships within multi-omic data. The transcriptome + covariates model achieved the highest AUROC (0.703) and AUPRC (0.622) among all tested models, showing a substantial improvement over its logistic regression counterpart. The proteome + covariates model also improved in performance (AUROC

= 0.680, AUPRC = 0.599), reinforcing the value of protein-level data in predictive modeling. The integration of both transcriptomic and proteomic data (EA and AA models) yielded AUROCs around 0.690 with comparable AUPRC values (∼0.613), indicating that multi-omic integration enhances predictive performance regardless of genetic similarity. The average, standard deviation, and confidence interval of all performance metrics across the 10 iterations of each model is shown in **Table 2**. Boxplots of performance for each tissue-specific model for elastic-net logistic regression are available in **Supplemental Figure S3A**, and for random forest in **Supplemental Figure S3B**. Across both modeling approaches, the F1-score and balanced accuracy metrics followed similar trends, with random forest models consistently achieving superior results, suggesting improved classification performance. These findings highlight the importance of multi-omics integration and non-linear modeling approaches for improving predictive accuracy in complex phenotypes.

### 3.4 Feature importance

To assess feature importance in the best performing RF model, we used the Gini index to quantify the relative contribution of each feature to classification performance. Feature importance across 10 iterations per tissue type was determined. Across the top 5 tissues, brain cortex, whole blood, brain frontal cortex BA9, brain nucleus accumbens basal ganglia, and brain substantia nigra, the importance of each feature was averaged, as seen in **Figure 3A**. Our analysis revealed that principal components (PCs) and age were among the most influential predictors. PC4 exhibited the highest importance (0.1387), followed by age (0.0761) and PC1 (0.0750). PC2 (0.0630) and PC5 (0.0414) also demonstrated moderate importance, indicating that genetic similarity and demographic factors played a substantial role in the model’s predictions.

**Figure 3.**
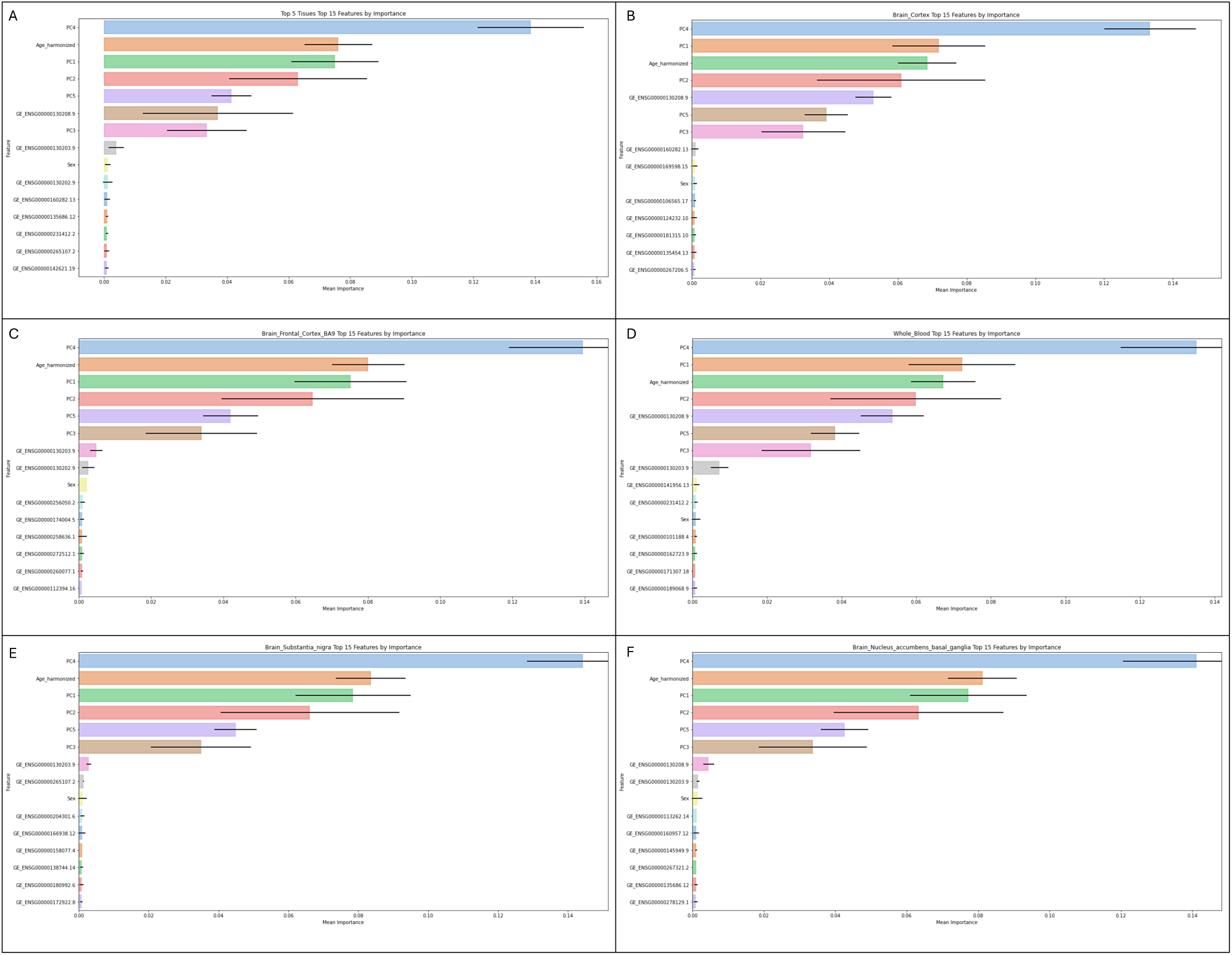
Feature importance rankings across top transcriptomic tissues. (A) Mean feature importance values across the top five tissues: brain cortex, whole blood, brain frontal cortex BA9, brain nucleus accumbens basal ganglia, and brain substantia nigra, highlighting consistently predictive genes. (B–F) Tissue-specific feature importance plots showing individual gene contributions to model performance within each tissue.

While gene expression (GE) features showed lower individual Gini importance scores compared to PCs, they remain biologically relevant. *APOC1* (ENSG00000130208.9) (0.0370) had an importance score comparable to PC5, suggesting that tissue-specific molecular variation contributes meaningfully to the model. Similarly, *APOE* (ENSG00000130203.9) (0.0040) and *NECTIN2* (ENSG00000130202.9) (0.0012) displayed non-negligible importance, though their relative contributions were lower. The comparatively modest importance of GE features may be partially attributed to their tissue specificity, as opposed to PCs, which were computed across individuals and therefore capture broader population-wide variance.

When tissue-specific contributions were individually assessed for feature importance (**Figure 3B-3F**), additional genes appeared significant, indicating the importance of tissue-specific variation. Across all 5 top tissues, PCs and age emerged as the dominant predictors, consistently ranking among the most important features. PC4 was the top predictor in every tissue, with the highest importance observed in the brain frontal cortex (0.1395) and slightly lower but comparable values in the brain cortex (0.1334) and whole blood (0.1351). This component was expected to be the most important as it separated the EUR genetic similarity group making up the greatest proportion of the samples from other populations, as shown in **Supplemental Figure S4**. Similarly, PC1, PC2, and PC5 maintained moderate importance across all tissues, suggesting that genetic similarity and demographic factors exert a broad influence on model predictions. The importance of age varied slightly, with the highest impact in the brain frontal cortex (0.0801), followed by the brain cortex (0.0686) and whole blood (0.0673). This suggests that age-related effects may be more pronounced in brain tissues compared to blood, which aligns with known age-related transcriptional changes in the central nervous system[51]. Tissue-specific feature importance for the remaining 12 tissues is shown in **Supplemental Figure S5A-L**.

GE feature importance varied significantly between tissues, highlighting the context-dependent role of molecular markers. *APOC1* (ENSG00000130208.9) was among the most important GE features in the brain cortex (0.0529) and whole blood (0.0536), but had lower importance in the brain nucleus accumbens (0.0045) and was absent from the top features in other brain regions. Similarly, *APOE* (ENSG00000130203.9) appeared in multiple tissues but with variable importance, ranging from 0.0073 in whole blood to 0.0016 in the brain nucleus accumbens. Brain substantia nigra prioritized a different set of GE features, with *GJA5* (ENSG00000265107.2) (0.0013) ranking among the top contributors.

## 4. Discussion

Our study identified a number of significant genomic, transcriptomic, and proteomic associations with Alzheimer’s Disease (AD) through multi-omics association studies. In addition to well-established loci within genes such as *APOE*. *APOC1*, *TOMM40*, *NECTIN2*, and *CR1*, we also uncovered several putatively novel signals which have been implicated in related biological processes linked to AD progression. Of the 104 genomic, 319 transcriptomic, and 17 proteomic associations, 122 unique genes and proteins were identified as statistically significant. Of these, 9 were independently validated in ADVP, providing additional support for the robustness of our association results. Many of the remaining associations were identified as contributors to AD pathology in literature searches and were statistically significant in the GWAS Catalog. These findings not only reinforce known genetic contributors to AD but also suggest new avenues for further biological investigation. For instance, *BLOC1S5-TXNDC5* encodes proteins involved in protein folding and the endoplasmic reticulum stress response. Notably, increased expression of *TXNDC5* has been linked to oxidative stress and neurodegeneration in AD[48]. *RUNX2*, while primarily known for its role in bone development, has also been suggested to have connections to neurodegenerative disease pathways[52]. *CBLC* functions in the ubiquitination and degradation of proteins—processes that, when disrupted, are known contributors to neurodegeneration[53].

From the TWAS, *PRDM16* has a known role in cellular differentiation in the developing brain, particularly in the production of mature neurons and their specific positions in the neocortex[54]. While it has not been directly linked to AD, it is associated with *PRDM10*, a gene previously listed in the ADVP[55]. *MRPL44*, which functions in mitochondrial protein synthesis, is also associated with *MRPL39* (an ADVP gene)[56]. Mitochondrial dysfunction is a consistent hallmark of neurodegenerative diseases, including AD[57]. *PPFIA4* has been linked to synaptic organization and neuronal signaling, processes known to be disrupted early in AD[58]. *CAPN2* encodes a calcium-dependent protease; dysregulation of calpain activity has been reported in neurodegenerative conditions, including AD[59]. *BTBD11* may be involved in transcriptional regulation and broader cellular processes via methylation[60]. *PALM3* plays a role in the Toll signaling pathway and in negative regulation of the cytokine-mediated signaling pathway[61]. *PALM2AKAP2*, a paralog of *PALM3*, is a known ADVP gene, and a recent preprint study identified associations between *PALM3* and neurodegenerative diseases[62]. Genes associated with mitochondrial function were also identified as significant in TWAS. *MTG2* contributes to mitochondrial maintenance, reinforcing the broader role of mitochondrial dysfunction in neurodegenerative conditions[56]. *SUCO*, although less characterized, is thought to be involved in maintaining cellular structure, which may impact neuronal integrity[63]. Other significantly expressed genes include *RASD1*, which participates in intracellular signaling pathways related to neuronal growth and synaptic plasticity, and *ZDHHC5*, a gene involved in protein palmitoylation and localized in neuronal structures[64,65]. *ZDHHC5* has been linked to neurological disorders and is associated with *ZDHHC20*, a gene recorded in the ADVP[65,66].

From the significant PWAS results, APP (β-amyloid precursor protein) was significantly associated with LOAD, consistent with its central role in β-amyloid plaque formation and disease progression[7]. Several additional proteins identified as significant showed functional relevance despite not being known AD risk genes. For example, PROS1 acts as a ligand for the TAM family of receptor tyrosine kinases, which help mediate anti-inflammatory responses and the phagocytic clearance of apoptotic cells and cellular debris, including potentially toxic aggregates like amyloid-β[67]. IL1RAP has been linked to increased amyloid accumulation independently of APOE ε4 status, suggesting alternative mechanisms of amyloid pathology[68]. ANXA7 is a calcium-dependent signaling protein in astrocytes; other annexins have been implicated in neuronal development and AD[69]. HADH, highly expressed in AD brains, binds Aβ and contributes to its cytotoxicity[70]. SERPINE1, encoding PAI-1, is associated with Aβ accumulation and broader neurodegenerative processes[71]. ST6GALNAC6, functionally related to the ADVP gene *ST6GAL2*, has been implicated in glycosylation changes observed in AD[32,72]. The results of the association studies implicate several potential biological mechanisms underlying AD risk that the gene-set enrichment analysis further highlight.

Pathway enrichment analysis of the genomic, transcriptomic, and proteomic association results offer complementary insights into AD biology, with each modality highlighting distinct aspects of disease etiology. The GWAS-based gene set enrichment identified robust associations with lipid metabolism, particularly plasma lipoprotein assembly, remodeling, and cholesterol efflux pathways. These findings are consistent with established roles of apolipoproteins, *APOE* and *APOC1*, in modulating amyloid deposition and neuronal integrity through cholesterol transport and lipid homeostasis[7]. The TWAS-based enrichment analysis did not yield statistically significant pathways after correction, likely due to the relatively small number of tissue-specific genes available for transcriptomic association testing. Nominal enrichments in transcriptional regulation and DNA repair pathways, such as RNA polymerase II preinitiation complex assembly, suggest effects of gene regulation and genome maintenance mechanisms that may be further revealed in larger AD datasets. Notably, the PWAS analysis revealed a strong signal for immune-related pathways, including interleukin signaling and inflammatory response regulation, implicating immune dysregulation in AD risk[7,73]. This immune enrichment was distinct from the lipid-related processes emphasized in GWAS, suggesting that the proteomic layer may capture systemic or downstream consequences of AD-related perturbations[73]. These results underscore the value of integrating multi-omics approaches to dissect the complex, multifactorial nature of AD, and point toward the convergence of lipid metabolism, transcriptional regulation, and immune signaling in shaping disease susceptibility.

Building on these molecular insights, our IRMs demonstrated substantial improvements over traditional prediction approaches. Notably, models incorporating imputed transcriptomic and proteomic expression features consistently outperformed baseline regression models of covariates, as well as PGS across nearly all evaluated metrics (AUROC, AUPRC, balanced accuracy). The PGS constructed using scores derived from IGAP, one of the largest and most robust GWAS efforts in AD, demonstrated modest improvements in AUROC relative to polygenic scores derived from internal ADSP splits. However, across other key metrics such as AUPRC, balanced accuracy, and F1 score, the IGAP-based model underperformed. Notably, even this best-case GWAS-driven approach failed to match the predictive performance of the IRMs. These findings underscore the limitations of relying solely on GWAS-derived polygenic scores, particularly when evaluating model utility across genetically-dissimilar populations. The superior performance of the IRMs highlights the added value of incorporating transcriptomic and proteomic information, which provide tissue- and context-specific biological signals absent in traditional PGS frameworks. Moreover, the random forest models further outperformed all linear models, reflecting the utility of non-linear methods in capturing complex, multi-omic relationships. Within the IRMs, principal components (PCs) emerged as dominant predictors, likely due to their role in capturing large-scale genetic similarity and population structure effects. GE features also contributed significantly, particularly in tissues such as whole blood and specific brain regions. Tissue-specific variability in GE feature importance suggests that expression regulation in distinct biological contexts meaningfully influences disease risk.

There are several limitations and opportunities for improvement in the development of these IRMs. GREx and GRPx inherently reflect predicted, rather than directly measured molecular phenotypes, based on individual-level genotype data. This was an intentional design choice to enable broader applicability across biobanks where transcriptomic or proteomic profiling may not be available, thereby leveraging scalable prediction models trained on external QTL reference panels to predict heritable risk of developing AD. They may be sensitive to imputation error, especially across different genetic similarity groups, as prediction accuracy is highly dependent on the ancestral composition and size of the reference QTL panels. In the future, this framework can be employed to integrate and assess sequenced genomic, transcriptomic and proteomic data rather than relying on imputed information, as such datasets become more readily available. Additionally, the tissue-specific nature of GE limits the completeness of prediction models, as not all relevant tissues (e.g., microglia or specific hippocampal subregions) were captured in the available resources. Environmental factors and cellular heterogeneity, which are not reflected in static genomic data, also introduce sources of unexplained variance. Future iterations of these models can account for additional contextual layers of regulation by incorporating additional relevant tissues, and integrating longitudinal multi-omics data to better model dynamic biological processes and tissue-specific effects more accurately. Moreover, although our models are adjusted for PCs, population stratification remains a potential confounder in genetic risk prediction, and future work should prioritize increasing sample size across populations.

This study demonstrates the clinical and biological value of integrating genetically-regulated gene and protein expression with genetic variants for AD risk prediction. Our IRM framework advances beyond traditional PGS approaches by incorporating biologically meaningful, tissue-specific regulatory information. The improved predictive performance, particularly with non-linear modeling, underscores the potential of multi-omics integration to enhance precision medicine strategies. Importantly, the generalizability of our framework offers a scalable pathway for expanding this approach to other complex traits and diseases, ultimately supporting the development of individualized risk prediction tools for clinical decision support.

## Data Availability

All summary-level results produced in the present study will be made available. All reference data used are available online through the 1000 Genomes Project, Genotype-Tissue Expression Project (GTEx v8) and Atherosclerosis Risk in Communities (ARIC) Study.

## 5. Acknowledgements

**ADSP Acknowledgement Statement**

Data for this study were prepared, archived, and distributed by the National Institute on Aging Alzheimer’s Disease Data Storage Site (NIAGADS) at the University of Pennsylvania (U24-AG041689), funded by the National Institute on Aging.

The Alzheimer’s Disease Sequencing Project (ADSP) is comprised of two Alzheimer’s Disease (AD) genetics consortia and three National Human Genome Research Institute (NHGRI) funded Large Scale Sequencing and Analysis Centers (LSAC). The two AD genetics consortia are the Alzheimer’s Disease Genetics Consortium (ADGC) funded by NIA (U01 AG032984), and the Cohorts for Heart and Aging Research in Genomic Epidemiology (CHARGE) funded by NIA (R01 AG033193), the National Heart, Lung, and Blood Institute (NHLBI), other National Institute of Health (NIH) institutes and other foreign governmental and non-governmental organizations. The Discovery Phase analysis of sequence data is supported through UF1AG047133 (to Drs. Schellenberg, Farrer, Pericak-Vance, Mayeux, and Haines); U01AG049505 to Dr. Seshadri; U01AG049506 to Dr. Boerwinkle; U01AG049507 to Dr. Wijsman; and U01AG049508 to Dr. Goate and the Discovery Extension Phase analysis is supported through U01AG052411 to Dr. Goate, U01AG052410 to Dr. Pericak-Vance and U01 AG052409 to Drs. Seshadri and Fornage.

Sequencing for the Follow Up Study (FUS) is supported through U01AG057659 (to Drs. PericakVance, Mayeux, and Vardarajan) and U01AG062943 (to Drs. Pericak-Vance and Mayeux). Data generation and harmonization in the Follow-up Phase is supported by U54AG052427 (to Drs. Schellenberg and Wang). The FUS Phase analysis of sequence data is supported through U01AG058589 (to Drs. Destefano, Boerwinkle, De Jager, Fornage, Seshadri, and Wijsman), U01AG058654 (to Drs. Haines, Bush, Farrer, Martin, and Pericak-Vance), U01AG058635 (to Dr. Goate), RF1AG058066 (to Drs. Haines, Pericak-Vance, and Scott), RF1AG057519 (to Drs. Farrer and Jun), R01AG048927 (to Dr. Farrer), and RF1AG054074 (to Drs. Pericak-Vance and Beecham).

The ADGC cohorts include: Adult Changes in Thought (ACT) (U01 AG006781, U19 AG066567), the Alzheimer’s Disease Research Centers (ADRC) (P30 AG062429, P30 AG066468, P30 AG062421, P30 AG066509, P30 AG066514, P30 AG066530, P30 AG066507, P30 AG066444, P30 AG066518, P30 AG066512, P30 AG066462, P30 AG072979, P30 AG072972, P30 AG072976, P30 AG072975, P30 AG072978, P30 AG072977, P30 AG066519, P30 AG062677, P30 AG079280, P30 AG062422, P30 AG066511, P30 AG072946, P30 AG062715, P30 AG072973, P30 AG066506, P30 AG066508, P30 AG066515, P30 AG072947, P30 AG072931, P30 AG066546, P20 AG068024, P20 AG068053, P20 AG068077, P20 AG068082, P30 AG072958, P30 AG072959), the Chicago Health and Aging Project (CHAP) (R01 AG11101, RC4 AG039085, K23 AG030944), Indiana Memory and Aging Study (IMAS) (R01 AG019771), Indianapolis Ibadan (R01 AG009956, P30 AG010133), the Memory and Aging Project (MAP) ( R01 AG17917), Mayo Clinic (MAYO) (R01 AG032990, U01 AG046139, R01 NS080820, RF1 AG051504, P50 AG016574), Mayo Parkinson’s Disease controls (NS039764, NS071674, 5RC2HG005605), University of Miami (R01 AG027944, R01 AG028786, R01 AG019085, IIRG09133827, A2011048), the Multi-Institutional Research in Alzheimer’s Genetic Epidemiology Study (MIRAGE) (R01 AG09029, R01 AG025259), the National Centralized Repository for Alzheimer’s Disease and Related Dementias (NCRAD) (U24 AG021886), the National Institute on Aging Late Onset Alzheimer’s Disease Family Study (NIA-LOAD) (U24 AG056270), the Religious Orders Study (ROS) (P30 AG10161, R01 AG15819), the Texas Alzheimer’s Research and Care Consortium (TARCC) (funded by the Darrell K Royal Texas Alzheimer’s Initiative), Vanderbilt University/Case Western Reserve University (VAN/CWRU) (R01 AG019757, R01 AG021547, R01 AG027944, R01 AG028786, P01 NS026630, and Alzheimer’s Association), the Washington Heights-Inwood Columbia Aging Project (WHICAP) (RF1 AG054023), the University of Washington Families (VA Research Merit Grant, NIA: P50AG005136, R01AG041797, NINDS: R01NS069719), the Columbia University Hispanic Estudio Familiar de Influencia Genetica de Alzheimer (EFIGA) (RF1 AG015473), the University of Toronto (UT) (funded by Wellcome Trust, Medical Research Council, Canadian Institutes of Health Research), and Genetic Differences (GD) (R01 AG007584). The CHARGE cohorts are supported in part by National Heart, Lung, and Blood Institute (NHLBI) infrastructure grant HL105756 (Psaty), RC2HL102419 (Boerwinkle) and the neurology working group is supported by the National Institute on Aging (NIA) R01 grant AG033193.

The CHARGE cohorts participating in the ADSP include the following: Austrian Stroke Prevention Study (ASPS), ASPS-Family study, and the Prospective Dementia Registry-Austria (ASPS/PRODEM-Aus), the Atherosclerosis Risk in Communities (ARIC) Study, the Cardiovascular Health Study (CHS), the Erasmus Rucphen Family Study (ERF), the Framingham Heart Study (FHS), and the Rotterdam Study (RS). ASPS is funded by the Austrian Science Fond (FWF) grant number P20545-P05 and P13180 and the Medical University of Graz. The ASPS-Fam is funded by the Austrian Science Fund (FWF) project I904), the EU Joint Programme – Neurodegenerative Disease Research (JPND) in frame of the BRIDGET project (Austria, Ministry of Science) and the Medical University of Graz and the Steiermärkische Krankenanstalten Gesellschaft. PRODEM-Austria is supported by the Austrian Research Promotion agency (FFG) (Project No. 827462) and by the Austrian National Bank (Anniversary Fund, project 15435. ARIC research is carried out as a collaborative study supported by NHLBI contracts (HHSN268201100005C, HHSN268201100006C, HHSN268201100007C, HHSN268201100008C, HHSN268201100009C, HHSN268201100010C, HHSN268201100011C, and HHSN268201100012C). Neurocognitive data in ARIC is collected by U01 2U01HL096812, 2U01HL096814, 2U01HL096899, 2U01HL096902, 2U01HL096917 from the NIH (NHLBI, NINDS, NIA and NIDCD), and with previous brain MRI examinations funded by R01-HL70825 from the NHLBI. CHS research was supported by contracts HHSN268201200036C, HHSN268200800007C, N01HC55222, N01HC85079, N01HC85080, N01HC85081, N01HC85082, N01HC85083, N01HC85086, and grants U01HL080295 and U01HL130114 from the NHLBI with additional contribution from the National Institute of Neurological Disorders and Stroke (NINDS). Additional support was provided by R01AG023629, R01AG15928, and R01AG20098 from the NIA. FHS research is supported by NHLBI contracts N01-HC-25195 and HHSN268201500001I. This study was also supported by additional grants from the NIA (R01s AG054076, AG049607 and AG033040 and NINDS (R01 NS017950). The ERF study as a part of EUROSPAN (European Special Populations Research Network) was supported by European Commission FP6 STRP grant number 018947 (LSHG-CT-2006-01947) and also received funding from the European Community’s Seventh Framework Programme (FP7/2007-2013)/grant agreement HEALTH-F4-2007-201413 by the European Commission under the programme “Quality of Life and Management of the Living Resources” of 5th Framework Programme (no. QLG2-CT-2002-01254). High-throughput analysis of the ERF data was supported by a joint grant from the Netherlands Organization for Scientific Research and the Russian Foundation for Basic Research (NWO-RFBR 047.017.043). The Rotterdam Study is funded by Erasmus Medical Center and Erasmus University, Rotterdam, the Netherlands Organization for Health Research and Development (ZonMw), the Research Institute for Diseases in the Elderly (RIDE), the Ministry of Education, Culture and Science, the Ministry for Health, Welfare and Sports, the European Commission (DG XII), and the municipality of Rotterdam. Genetic data sets are also supported by the Netherlands Organization of Scientific Research NWO Investments (175.010.2005.011, 911-03-012), the Genetic Laboratory of the Department of Internal Medicine, Erasmus MC, the Research Institute for Diseases in the Elderly (014-93-015; RIDE2), and the Netherlands Genomics Initiative (NGI)/Netherlands Organization for Scientific Research (NWO) Netherlands Consortium for Healthy Aging (NCHA), project 050-060-810. All studies are grateful to their participants, faculty and staff. The content of these manuscripts is solely the responsibility of the authors and does not necessarily represent the official views of the National Institutes of Health or the U.S. Department of Health and Human Services.

The FUS cohorts include: the Alzheimer’s Disease Research Centers (ADRC) (P30 AG062429, P30 AG066468, P30 AG062421, P30 AG066509, P30 AG066514, P30 AG066530, P30 AG066507, P30 AG066444, P30 AG066518, P30 AG066512, P30 AG066462, P30 AG072979, P30 AG072972, P30 AG072976, P30 AG072975, P30 AG072978, P30 AG072977, P30 AG066519, P30 AG062677, P30 AG079280, P30 AG062422, P30 AG066511, P30 AG072946, P30 AG062715, P30 AG072973, P30 AG066506, P30 AG066508, P30 AG066515, P30 AG072947, P30 AG072931, P30 AG066546, P20 AG068024, P20 AG068053, P20 AG068077, P20 AG068082, P30 AG072958, P30 AG072959), Alzheimer’s Disease Neuroimaging Initiative (ADNI) (U19AG024904), Amish Protective Variant Study (RF1AG058066), Cache County Study (R01AG11380, R01AG031272, R01AG21136, RF1AG054052), Case Western Reserve University Brain Bank (CWRUBB) (P50AG008012), Case Western Reserve University Rapid Decline (CWRURD) (RF1AG058267, NU38CK000480), CubanAmerican Alzheimer’s Disease Initiative (CuAADI) (3U01AG052410), Estudio Familiar de Influencia Genetica en Alzheimer (EFIGA) (5R37AG015473, RF1AG015473, R56AG051876), Genetic and Environmental Risk Factors for Alzheimer Disease Among African Americans Study (GenerAAtions) (2R01AG09029, R01AG025259, 2R01AG048927), Gwangju Alzheimer and Related Dementias Study (GARD) (U01AG062602), Hillblom Aging Network (2014-A-004-NET, R01AG032289, R01AG048234), Hussman Institute for Human Genomics Brain Bank (HIHGBB) (R01AG027944, Alzheimer’s Association “Identification of Rare Variants in Alzheimer Disease”), Ibadan Study of Aging (IBADAN) (5R01AG009956), Longevity Genes Project (LGP) and LonGenity (R01AG042188, R01AG044829, R01AG046949, R01AG057909, R01AG061155, P30AG038072), Mexican Health and Aging Study (MHAS) (R01AG018016), Multi-Institutional Research in Alzheimer’s Genetic Epidemiology (MIRAGE) (2R01AG09029, R01AG025259, 2R01AG048927), Northern Manhattan Study (NOMAS) (R01NS29993), Peru Alzheimer’s Disease Initiative (PeADI) (RF1AG054074), Puerto Rican 1066 (PR1066) (Wellcome Trust (GR066133/GR080002), European Research Council (340755)), Puerto Rican Alzheimer Disease Initiative (PRADI) (RF1AG054074), Reasons for Geographic and Racial Differences in Stroke (REGARDS) (U01NS041588), Research in African American Alzheimer Disease Initiative (REAAADI) (U01AG052410), the Religious Orders Study (ROS) (P30 AG10161, P30 AG72975, R01 AG15819, R01 AG42210), the RUSH Memory and Aging Project (MAP) (R01 AG017917, R01 AG42210Stanford Extreme Phenotypes in AD (R01AG060747), University of Miami Brain Endowment Bank (MBB), University of Miami/Case Western/North Carolina A&T African American (UM/CASE/NCAT) (U01AG052410, R01AG028786), Wisconsin Registry for Alzheimer’s Prevention (WRAP) (R01AG027161 and R01AG054047), Mexico-Southern California Autosomal Dominant Alzheimer’s Disease Consortium (R01AG069013), Center for Cognitive Neuroscience and Aging (R01AG047649), and the A4 Study (R01AG063689, U19AG010483 and U24AG057437).

The four LSACs are: the Human Genome Sequencing Center at the Baylor College of Medicine (U54 HG003273), the Broad Institute Genome Center (U54HG003067), The American Genome Center at the Uniformed Services University of the Health Sciences (U01AG057659), and the Washington University Genome Institute (U54HG003079). Genotyping and sequencing for the ADSP FUS is also conducted at John P. Hussman Institute for Human Genomics (HIHG) Center for Genome Technology (CGT).

Biological samples and associated phenotypic data used in primary data analyses were stored at Study Investigators institutions, and at the National Centralized Repository for Alzheimer’s Disease and Related Dementias (NCRAD, U24AG021886) at Indiana University funded by NIA. Associated Phenotypic Data used in primary and secondary data analyses were provided by Study Investigators, the NIA funded Alzheimer’s Disease Centers (ADCs), and the National Alzheimer’s Coordinating Center (NACC, U24AG072122) and the National Institute on Aging Genetics of Alzheimer’s Disease Data Storage Site (NIAGADS, U24AG041689) at the University of Pennsylvania, funded by NIA. Harmonized phenotypes were provided by the ADSP Phenotype Harmonization Consortium (ADSP-PHC), funded by NIA (U24 AG074855, U01 AG068057 and R01 AG059716) and Ultrascale Machine Learning to Empower Discovery in Alzheimer’s Disease Biobanks (AI4AD, U01 AG068057). This research was supported in part by the Intramural Research Program of the National Institutes of health, National Library of Medicine. Contributors to the Genetic Analysis Data included Study Investigators on projects that were individually funded by NIA, and other NIH institutes, and by private U.S. organizations, or foreign governmental or nongovernmental organizations.

The ADSP Phenotype Harmonization Consortium (ADSP-PHC) is funded by NIA (U24 AG074855, U01 AG068057 and R01 AG059716). The harmonized cohorts within the ADSP-PHC include: the Anti-Amyloid Treatment in Asymptomatic Alzheimer’s study (A4 Study), a secondary prevention trial in preclinical Alzheimer’s disease, aiming to slow cognitive decline associated with brain amyloid accumulation in clinically normal older individuals. The A4 Study is funded by a public-private-philanthropic partnership, including funding from the National Institutes of Health-National Institute on Aging, Eli Lilly and Company, Alzheimer’s Association, Accelerating Medicines Partnership, GHR Foundation, an anonymous foundation and additional private donors, with in-kind support from Avid and Cogstate. The companion observational Longitudinal Evaluation of Amyloid Risk and Neurodegeneration (LEARN) Study is funded by the Alzheimer’s Association and GHR Foundation. The A4 and LEARN Studies are led by Dr. Reisa Sperling at Brigham and Women’s Hospital, Harvard Medical School and Dr. Paul Aisen at the Alzheimer’s Therapeutic Research Institute (ATRI), University of Southern California. The A4 and LEARN Studies are coordinated by ATRI at the University of Southern California, and the data are made available through the Laboratory for Neuro Imaging at the University of Southern California. The participants screening for the A4 Study provided permission to share their de-identified data in order to advance the quest to find a successful treatment for Alzheimer’s disease. We would like to acknowledge the dedication of all the participants, the site personnel, and all of the partnership team members who continue to make the A4 and LEARN Studies possible. The complete A4 Study Team list is available on: a4study.org/a4-study-team.; the Adult Changes in Thought study (ACT), U01 AG006781, U19 AG066567; Alzheimer’s Disease Neuroimaging Initiative (ADNI): Data collection and sharing for this project was funded by the Alzheimer’s Disease Neuroimaging Initiative (ADNI) (National Institutes of Health Grant U01 AG024904) and DOD ADNI (Department of Defense award number W81XWH-12-2-0012). ADNI is funded by the National Institute on Aging, the National Institute of Biomedical Imaging and Bioengineering, and through generous contributions from the following: AbbVie, Alzheimer’s Association; Alzheimer’s Drug Discovery Foundation; Araclon Biotech; BioClinica, Inc.; Biogen; Bristol-Myers Squibb Company; CereSpir, Inc.; Cogstate; Eisai Inc.; Elan Pharmaceuticals, Inc.; Eli Lilly and Company; EuroImmun; F. Hoffmann-La Roche Ltd and its affiliated company Genentech, Inc.; Fujirebio; GE Healthcare; IXICO Ltd.;Janssen Alzheimer Immunotherapy Research & Development, LLC.; Johnson & Johnson Pharmaceutical Research & Development LLC.; Lumosity; Lundbeck; Merck & Co., Inc.;Meso Scale Diagnostics, LLC.; NeuroRx Research; Neurotrack Technologies; Novartis Pharmaceuticals Corporation; Pfizer Inc.; Piramal Imaging; Servier; Takeda Pharmaceutical Company; and Transition Therapeutics. The Canadian Institutes of Health Research is providing funds to support ADNI clinical sites in Canada. Private sector contributions are facilitated by the Foundation for the National Institutes of Health (www.fnih.org). The grantee organization is the Northern California Institute for Research and Education, and the study is coordinated by the Alzheimer’s Therapeutic Research Institute at the University of Southern California. ADNI data are disseminated by the Laboratory for Neuro Imaging at the University of Southern California; Estudio Familiar de Influencia Genetica en Alzheimer (EFIGA): 5R37AG015473, RF1AG015473, R56AG051876; the Health & Aging Brain Study – Health Disparities (HABS-HD), supported by the National Institute on Aging of the National Institutes of Health under Award Numbers R01AG054073, R01AG058533, R01AG070862, P41EB015922, and U19AG078109; the Korean Brain Aging Study for the Early Diagnosis and Prediction of Alzheimer’s disease (KBASE), which was supported by a grant from Ministry of Science, ICT and Future Planning (Grant No: NRF-2014M3C7A1046042); Memory & Aging Project at Knight Alzheimer’s Disease Research Center (MAP at Knight ADRC): The Memory and Aging Project at the Knight-ADRC (Knight-ADRC). This work was supported by the National Institutes of Health (NIH) grants R01AG064614, R01AG044546, RF1AG053303, RF1AG058501, U01AG058922 and R01AG064877 to Carlos Cruchaga. The recruitment and clinical characterization of research participants at Washington University was supported by NIH grants P30AG066444, P01AG03991, and P01AG026276. Data collection and sharing for this project was supported by NIH grants RF1AG054080, P30AG066462, R01AG064614 and U01AG052410. We thank the contributors who collected samples used in this study, as well as patients and their families, whose help and participation made this work possible. This work was supported by access to equipment made possible by the Hope Center for Neurological Disorders, the Neurogenomics and Informatics Center (NGI: https://neurogenomics.wustl.edu/) and the Departments of Neurology and Psychiatry at Washington University School of Medicine; National Alzheimer’s Coordinating Center (NACC): The NACC database is funded by NIA/NIH Grant U24 AG072122. SCAN is a multi-institutional project that was funded as a U24 grant (AG067418) by the National Institute on Aging in May 2020. Data collected by SCAN and shared by NACC are contributed by the NIA-funded ADRCs as follows: P30 AG062429 (PI James Brewer, MD, PhD), P30 AG066468 (PI Oscar Lopez, MD), P30 AG062421 (PI Bradley Hyman, MD, PhD), P30 AG066509 (PI Thomas Grabowski, MD), P30 AG066514 (PI Mary Sano, PhD), P30 AG066530 (PI Helena Chui, MD), P30 AG066507 (PI Marilyn Albert, PhD), P30 AG066444 (PI John Morris, MD), P30 AG066518 (PI Jeffrey Kaye, MD), P30 AG066512 (PI Thomas Wisniewski, MD), P30 AG066462 (PI Scott Small, MD), P30 AG072979 (PI David Wolk, MD), P30 AG072972 (PI Charles DeCarli, MD), P30 AG072976 (PI Andrew Saykin, PsyD), P30 AG072975 (PI David Bennett, MD), P30 AG072978 (PI Neil Kowall, MD), P30 AG072977 (PI Robert Vassar, PhD), P30 AG066519 (PI Frank LaFerla, PhD), P30 AG062677 (PI Ronald Petersen, MD, PhD), P30 AG079280 (PI Eric Reiman, MD), P30 AG062422 (PI Gil Rabinovici, MD), P30 AG066511 (PI Allan Levey, MD, PhD), P30 AG072946 (PI Linda Van Eldik, PhD), P30 AG062715 (PI Sanjay Asthana, MD, FRCP), P30 AG072973 (PI Russell Swerdlow, MD), P30 AG066506 (PI Todd Golde, MD, PhD), P30 AG066508 (PI Stephen Strittmatter, MD, PhD), P30 AG066515 (PI Victor Henderson, MD, MS), P30 AG072947 (PI Suzanne Craft, PhD), P30 AG072931 (PI Henry Paulson, MD, PhD), P30 AG066546 (PI Sudha Seshadri, MD), P20 AG068024 (PI Erik Roberson, MD, PhD), P20 AG068053 (PI Justin Miller, PhD), P20 AG068077 (PI Gary Rosenberg, MD), P20 AG068082 (PI Angela Jefferson, PhD), P30 AG072958 (PI Heather Whitson, MD), P30 AG072959 (PI James Leverenz, MD); National Institute on Aging Alzheimer’s Disease Family Based Study (NIA-AD FBS): U24 AG056270; Religious Orders Study (ROS): P30AG10161,R01AG15819, R01AG42210; Memory and Aging Project (MAP - Rush): R01AG017917, R01AG42210; Minority Aging Research Study (MARS): R01AG22018, R01AG42210; the Texas Alzheimer’s Research and Care Consortium (TARCC), funded by the Darrell K Royal Texas Alzheimer’s Initiative, directed by the Texas Council on Alzheimer’s Disease and Related Disorders; Washington Heights/Inwood Columbia Aging Project (WHICAP): RF1 AG054023;and Wisconsin Registry for Alzheimer’s Prevention (WRAP): R01AG027161 and R01AG054047. Additional acknowledgments include the National Institute on Aging Genetics of Alzheimer’s Disease Data Storage Site (NIAGADS, U24AG041689) at the University of Pennsylvania, funded by NIA.

## Consent Statement

All subjects were consented according to ADSP procedures, as described in the ADSP Acknowledgement Statement, prior to data acquisition and release.

## Conflicts

The authors declare that they have no competing interests and nothing to disclose.

## Author Contributions

RV: Conceptualization, data curation, analysis, methodology, software, visualization, writing - original draft, writing - review and editing.

KMC: Data curation, analysis, writing - review and editing.

YB: Data curation, analysis, writing - review and editing.

AKM: Data curation, analysis, writing - review and editing.

RK: Data curation, analysis, writing - review and editing.

JHM: Funding acquisition, supervision, writing - review and editing

LS: Funding acquisition, supervision, writing - review and editing

DK: Funding acquisition, supervision, writing - review and editing

MDR: Conceptualization, data curation, funding acquisition, resources, supervision, writing - review and editing.

## Funding Sources

This work was supported by the National Institutes of Health (HL-169458, U01 AG066833, P30AG073105). RK was partially supported by the Training Program in Computational Genomics grant from the National Human Genome Research Institute to the University of Pennsylvania (T32HG000046). MDR, JHM, and LS were supported by U01 AG066833. LS and DK were additionally supported by U19 AG074879. All funding bodies had no role in the analysis or interpretation of data in this study.

## Notes

### Competing Interest Statement

The authors have declared no competing interest.

### Author Declarations

All individual level and summary-level ADSP data are accessible through NIAGADS. All individual level data has been de-identified and NIAGADS users are prohibited from trying to reidentify individuals. Restricted-access data through the NIAGADS Data Sharing Service require an approved data access request to ensure compliance with privacy and ethical guidelines and in accordance with the terms outlined by the submitting Institutional Review Boards (IRBs) and the consent provided by research participants. Reference eQTL and pQTL data models from GTEx and ARIC are available through PredictDB.

