## Supplemental Figures 1-5 for "Integrative multi-omics approaches identify molecular pathways and improve Alzheimer’s Disease risk prediction"

**Supplemental Figure S1.** ADSP PCA Plots. (A) Scree plot for the PCA indicating that 3-4 PCs captures 80% of the cumulative variance. (B) Scatterplot showing projection of the ADSP population PCs against the 1000Genomes PCs.

**Supplemental Figure S2.** Manhattan plots and QQ plots of the GWAS results. (A) Manhattan plot of GWAS results. (B) QQ plots of GWAS with APOE included. (C) QQ plots of GWAS without APOE region.

**Supplemental Figure S3**. Tissue-specific IRM model performance plots, depicting AUROC, AUPRC, F1 score, and balanced accuracy. (A) Boxplots of elastic-net logistic regression models for each transcriptomic and proteomic feature set. (B) Boxplots of random forest models for each transcriptomic and proteomic feature set.

**Supplemental Figure S4.** PC pair plots showing projections of the ADSP population for PCs 1-5 against genetically inferred ancestry (GIA). PC4 separates the EUR genetic similarity group from the other populations.

**Supplemental Figure S5.** Tissue-specific feature importance for the remaining 12 tissues. (A-L) Gini index-determined features for brain spinal cord cervical c-1, colon sigmoid, liver, vagina, brain anterior cingulate cortex BA24, brain hypothalamus, brain caudate basal ganglia, brain cerebellar hemisphere, brain putamen basal ganglia, brain cerebellum, brain amygdala, and brain hippocampus.

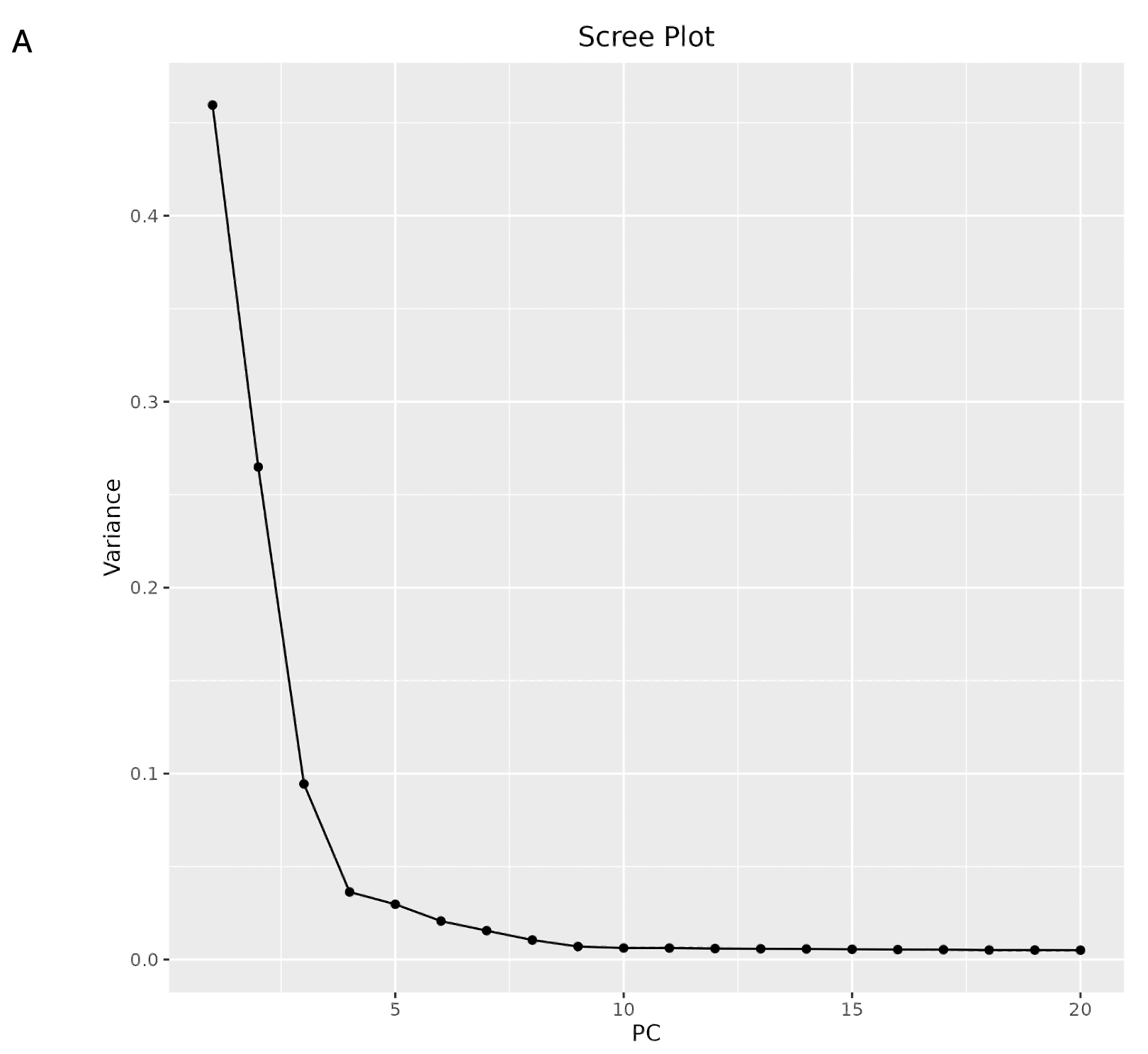

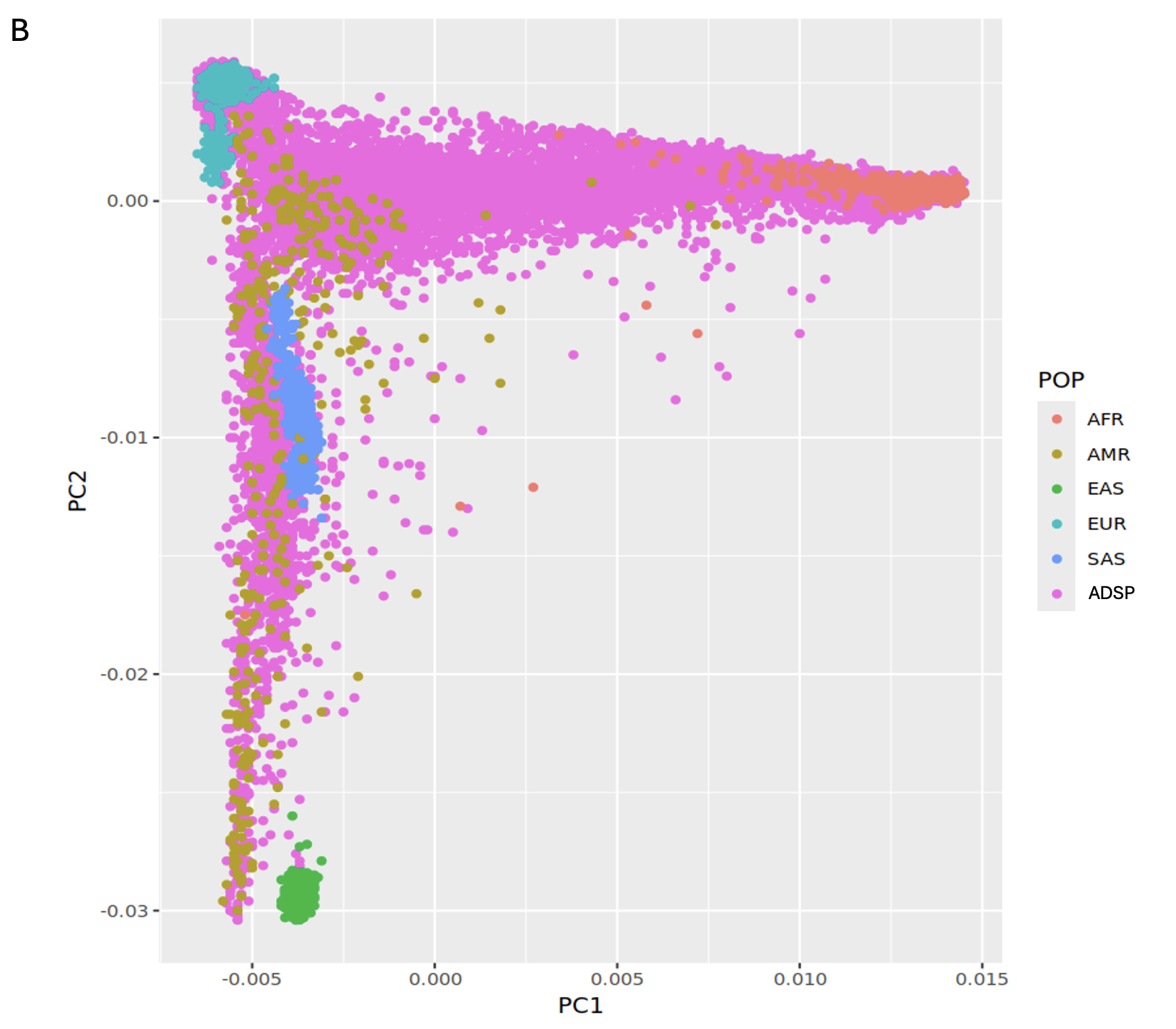

**Supplemental Figure S1.** ADSP PCA Plots. (A) Scree plot for the PCA indicating that 3-4 PCs captures 80% of the cumulative variance. (B) Scatterplot showing projection of the ADSP population PCs against the 1000Genomes PCs.

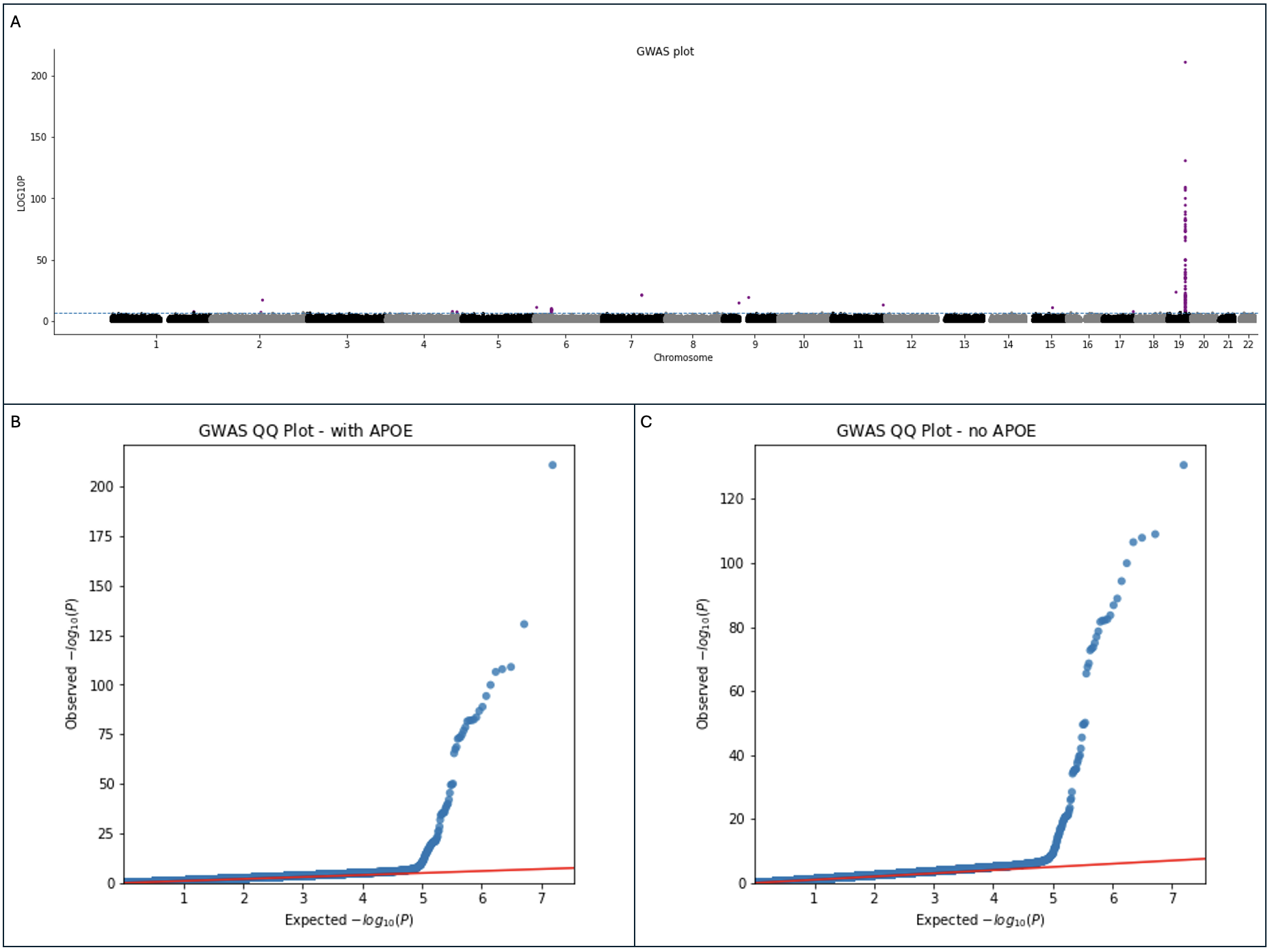

**Supplemental Figure S2.** Manhattan plots and QQ plots of the GWAS results. (A) Manhattan plot of GWAS results. (B) QQ plots of GWAS with APOE included. (C) QQ plots of GWAS without APOE region.

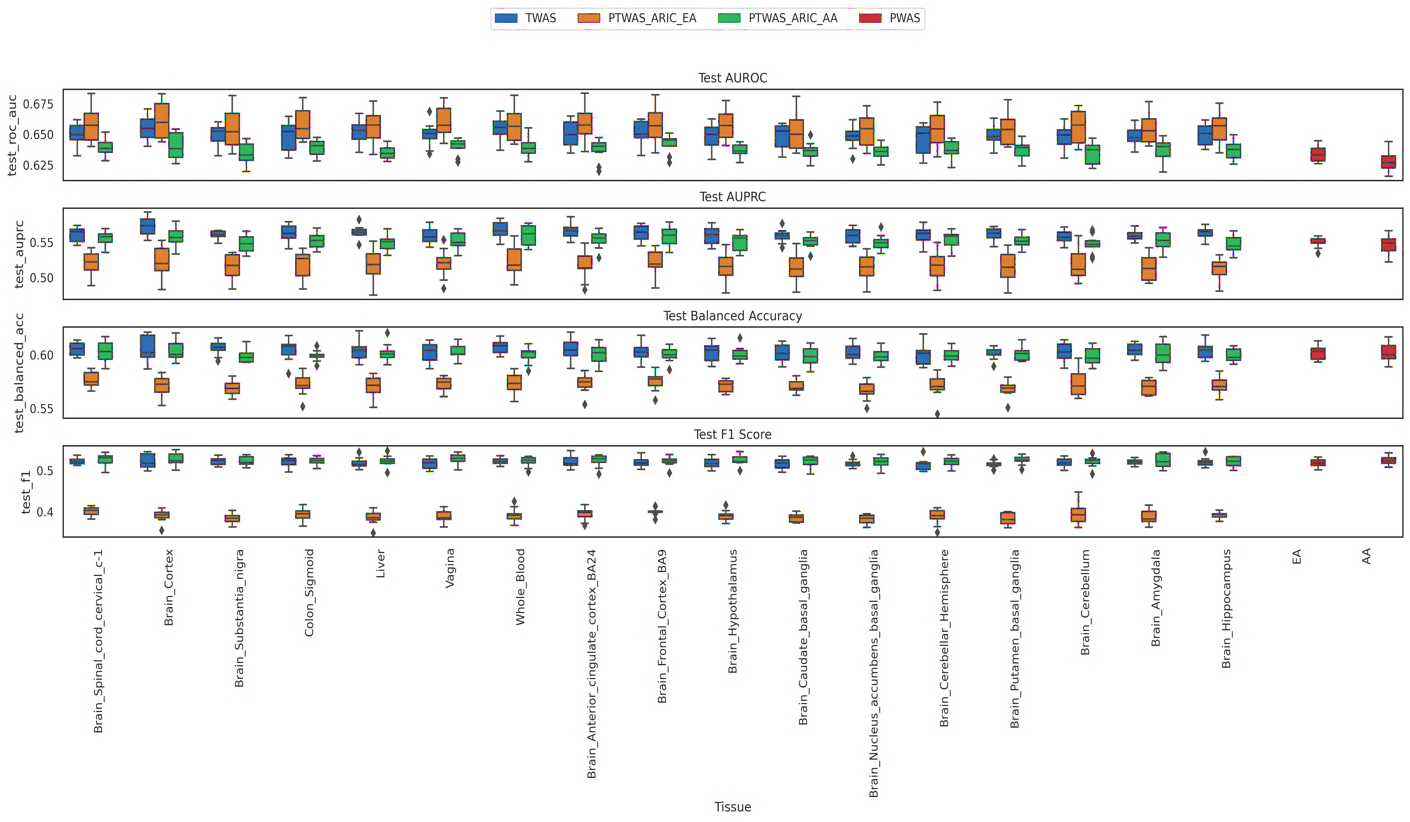
A

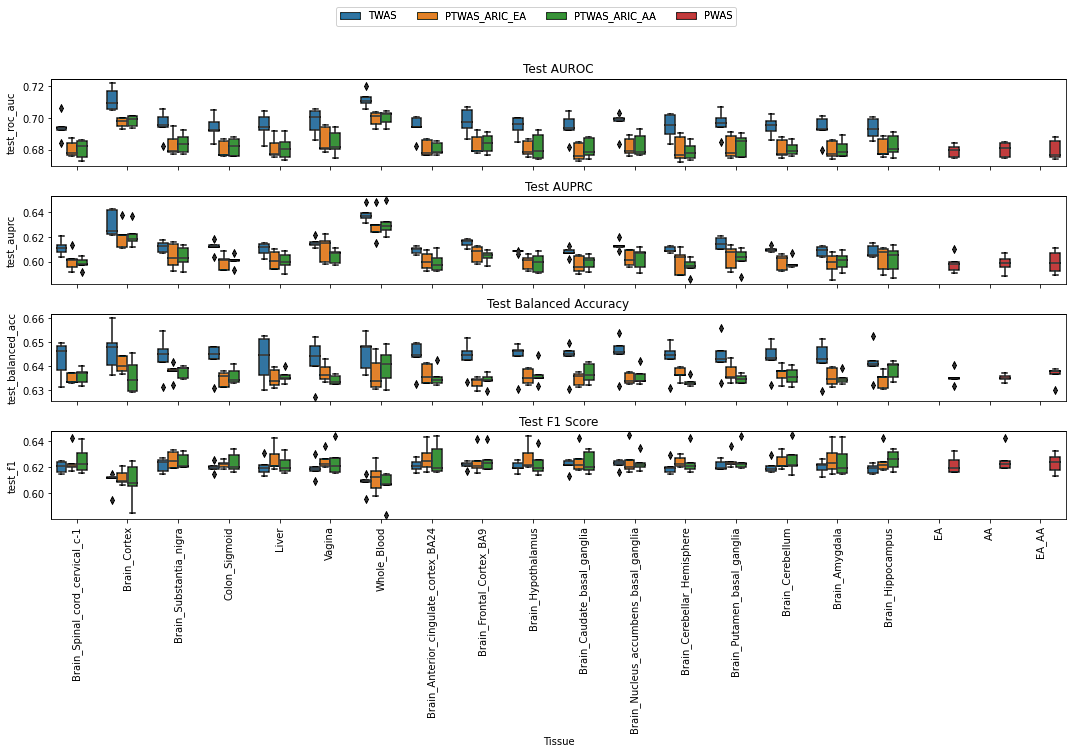

B

**Supplemental Figure S3**. Tissue-specific IRM model performance plots, depicting AUROC, AUPRC, F1 score, and balanced accuracy. (A) Boxplots of elastic-net logistic regression models for each transcriptomic and proteomic feature set. (B) Boxplots of random forest models for each transcriptomic and proteomic feature set.

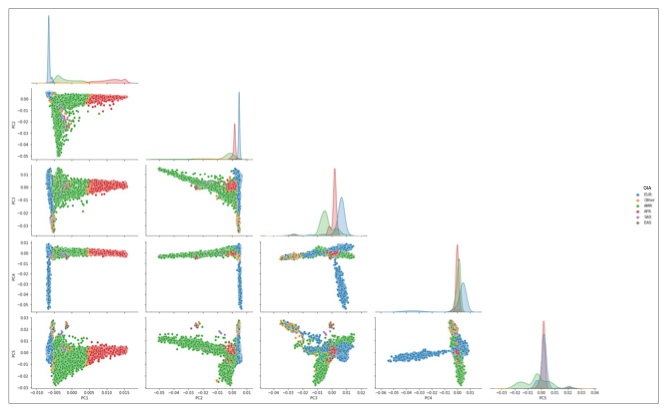

**Supplemental Figure S4.** PC pair plot showing projections of the ADSP participants for PCs 1-5 against genetically inferred ancestry (GIA). PC4 separates the EUR genetic similarity group from the other populations.

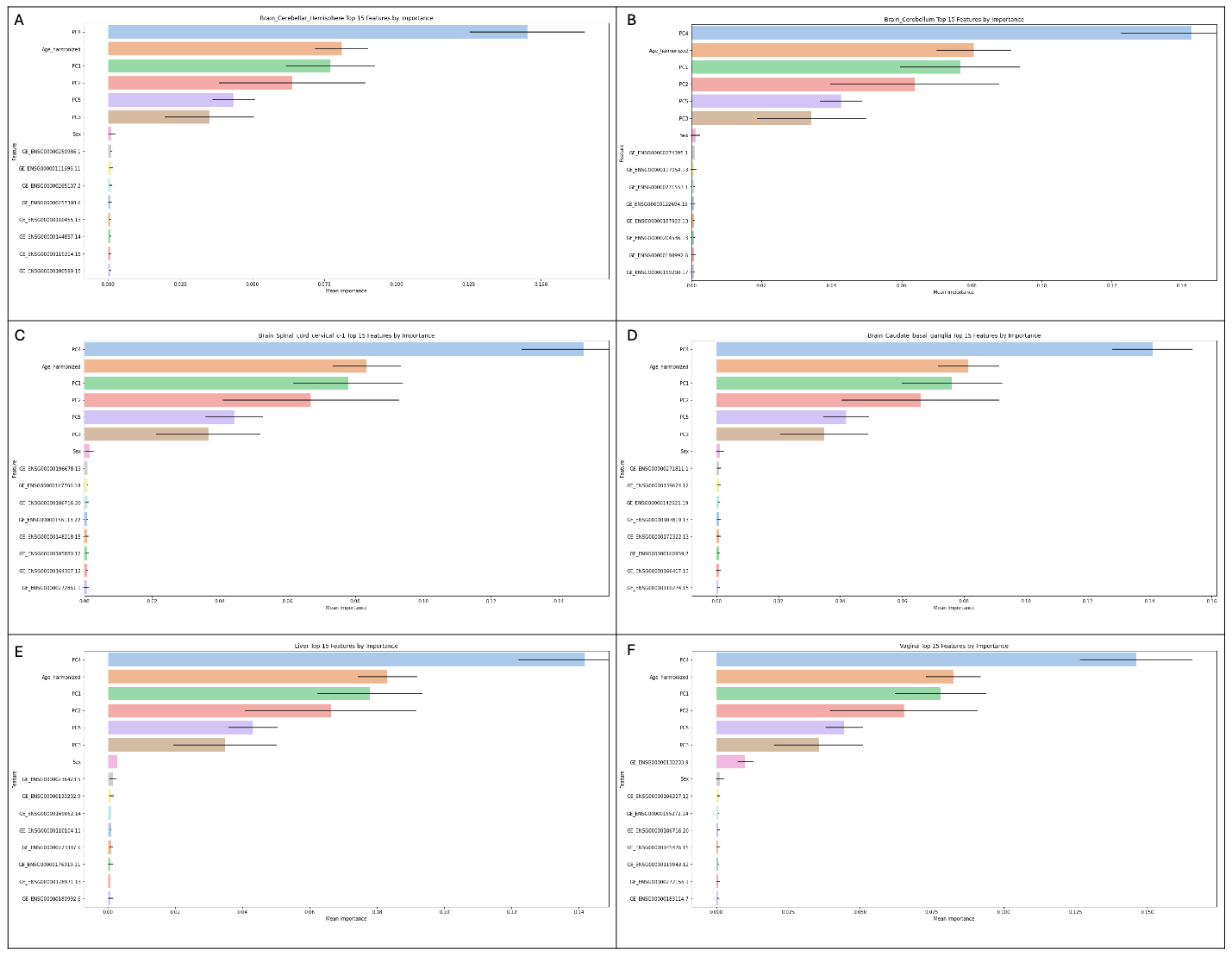

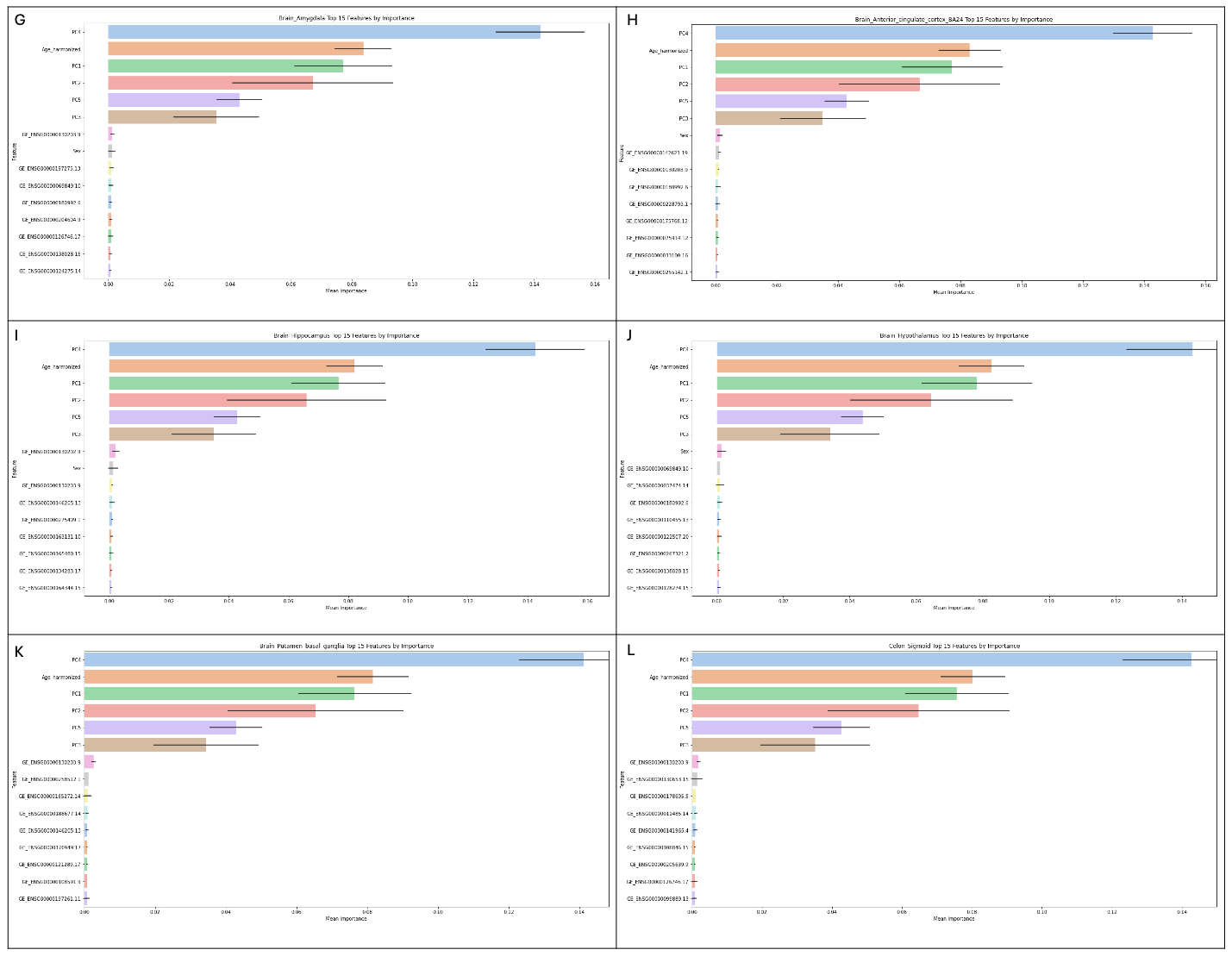

**Supplemental Figure S5.** Tissue-specific feature importance for the remaining 12 tissues. (A-L) Gini index-determined features for brain spinal cord cervical c-1, colon sigmoid, liver, vagina, brain anterior cingulate cortex BA24, brain hypothalamus, brain caudate basal ganglia, brain cerebellar hemisphere, brain putamen basal ganglia, brain cerebellum, brain amygdala, and brain hippocampus.
